# A systematic review to explore factors affecting participation in health research trials amongst underrepresented socio-economically disadvantaged populations in the UK and Ireland

**DOI:** 10.1101/2024.10.04.24313099

**Authors:** Jakki Brandon, Anne Marie Bagnall

## Abstract

**Background:** Those with the greatest health inequalities and highest burden of poor-health have the lowest representation in health research. Meaning that diseases which are known to have higher prevalence in disadvantaged populations are being studied in healthier populations, so findings will be inappropriately applied to those that face the greatest challenges to their health. This systematic review is concerned with those defined as socioeconomic-deprived, gypsy/traveller/Roma communities, and homeless people.

**Objectives:** To understand the barriers or facilitators to research participation in specific underserved groups to inform how equality, diversity and inclusion in research trials can be improved.

**Methods:** A systematic review of etiology of quantitative and qualitative primary research conducted in the UK and Ireland in the previous 10 years, to produce a narrative synthesis of the themes identified.

**Results:** Key themes of barriers and facilitators were identified across the groups. All facilitators reported were common to all the groups. The value of peer-researchers, gatekeepers, community relationships and participatory approaches was a prominent feature within the facilitators. Reporting of barriers overall was lower, with fewer commonalities seen across the groups. The SE deprived group had the fewest barriers reported.

**Conclusion:** The lack of identified barriers suggests that more work should be done to learn what these may be for specific groups, particularly in the SE deprived group. It seems likely that barriers exist in this group. The fact that these were not reported may indicate lack of reach to the most deprived sections of this population. Therefore, more research needs to be done to fully explore the barriers in the most deprived groups. By developing engagement approaches in a way which is tailored to separate underserved groups this will address barriers and help to shape the future of research.

## Background

There is burgeoning interest and increasing exploration of factors around underrepresentation in health research. This was accelerated to the fore during the Covid-19 pandemic when it was highlighted that there was disproportionately low representation in research trials from some population groups (Ekezie et al, 2021). A focus was placed upon black, Asian, minority ethnic (BAME) groups (Witham et al, 2020a). However, it became evident that this extended to other underserved populations, all of whom were known to carry a greater burden of incidence and effects of the disease (Ekezie et al, 2021).

Exploring participation in Covid-19 vaccines trials reflects some of the barriers and facilitators for underrepresented populations and shows some extent of the commonalities and divergences, not only in BAME populations but also other vulnerable underrepresented populations, including homeless, those with mental health difficulties and the gypsy/traveller/Roma community (Ekezie, 2021). Across diverse underrepresented groups Ekezie et al (2021) reported comparable themes of mistrust, misinformation, and apprehension, but endorse that different hesitancies between groups are considered carefully and distinctly so that approaches to reduce address barriers can be adapted accordingly.

Those with the greatest health inequalities and highest burden of poor health from long-term physical health conditions and mental ill-health are amongst the most underserved in health research (NIHR, 2021). This is estimated to be to such an extent that across 15 research networks approximately 12% of the research activity would need to be redistributed to effectively represent the underserved populations (Bower et al, 2020). A current, high-profile example of this is within dementia research. The NIHR have identified the need for greater research into dementia as high priority. People from BAME backgrounds have a greater propensity to dementia than the White British population, however, little is known about the association between ethnicity and dementia therefore screening and preventive strategies may not reach BAME populations (Tsamakis et al, 2021).

Whilst in no way diminishing the valid attention to the paucity of research in BAME groups, action to increase research participation in this population appears to outweigh that of other social economically vulnerable populations. Challenges around equality, diversity and inclusion (EDI) extends beyond the ethnicity of populations; BAME recruitment is relatively higher in less deprived areas and smaller cities as these areas tend to have lower economic deprivation and more educated BAME populations (Sethi et al, 2021).

Focussing specifically upon societal groups which suffer higher disease burdens should enable greater understanding of barriers or facilitators to research participation which may be unique to them, thus aiming to expand knowledge and evidence of disease incidence and processes in those underserved groups (Ekezie et al, 2021). Participation in research not only provides positive outcomes for the wider populations, but also improves health outcomes and satisfaction levels for individual patients (Krzyzanowska et al, 2011). Therefore, inadequate EDI for underrepresented individuals presents ethical issues of them having poorer health outcomes (Erves at al, 2017).

Underrepresented groups have historically been referred to as ‘hard-to-reach’, which creates the perception of the group being the problem (Bonevski et al, 2014). The focus is moving away from blaming the group, towards acknowledging the importance and the value of EDI in research participation. This should enable the benefits of clinical research participation for individuals from specific underrepresented groups and enable more equal representation across societies and populations (Bower et al, 2020).

### Rationale for the study

A 5-year analysis of participant data within NIHR funded clinical trials found that societal and population groups with the highest disease burden have the lowest representation in research (NIHR, 2021). This means that diseases which are known to have higher prevalence in deprived communities are being studied in healthier populations. This creates the risk of findings being inappropriately generalized to those communities that face greater challenges to their health. To endeavour to address this the NIHR formulated a 5-year plan to improve EDI in clinical research (NIHR, 2022a).

The author’s job role as clinical research nurse working in clinical trials, including vaccine research trials and early phase oncology trials has driven the interest to understand factors which may influence participation in clinical research. The author has been aware of local research initiatives within their own workplace to reach out to and improve engagement with BAME communities based around the NIHR research ready communities and community champion initiatives (NIHR, 2022b). The author is not aware of any equivalent initiatives within their workplace aimed at SE challenged groups and has seen first-hand the absences in research participation across many societal groups. The reason for conducting this systematic review (SR) is to draw together studies over the past 10 years exploring research participation in SE challenged groups in health research, in the UK and Ireland. These will be SE deprived, gypsy/traveller/Roma communities, and homeless people. It is envisaged that this will assist and inform the author, and the wider research community, as to how the approaches already seen within their workplace for engaging BAME populations may be applied to these chosen population groups.

### Aim of the study

This study aims to gain deeper understanding of how and why socioeconomic disadvantage and health inequalities may influence the likelihood of participating in health research.

### Objectives of the study

To perform a SR of etiology of primary research studies in the UK and Ireland between 2013-2023 to explore contributory factors behind underrepresentation in health research.

To understand barriers or facilitators to research participation unique to specific underserved groups to inform how EDI in research trials can be improved for SE deprived groups, GTRC and homeless people.

It is anticipated that this information will prove valuable in the field of patient and public engagement and involvement (PPIE) and will provide direction for PPIE with populations who experience high levels of health inequalities, disease burden and socio-economic challenges to improve research participation in underrepresented groups.

#### Literature overview

This literature overview uses a ‘who, what and why’ approach to explore and present a collection of relevant research and published literature.

### Who are the populations and what are their predominant health burdens and inequalities

#### Gypsy/traveller/Roma communities

The Gypsy/traveller/Roma community (GTRC) have long-since been one of the most excluded (Brown and Scullion, 2009), they continue to remain marginalized and experience severe health inequalities (Van Cleemput, 2018). The GTRC have poorer health status and disease outcomes across all disease groups and dramatically lower life expectancy compared to the wider population (Parry et al, 2007). Historically, lack of census data made it difficult to quantify, however more recently it is estimated that life expectancy may be as much as 28 years lower than the national average (Van Cleemput, 2018). The most evident health problems are mental-health difficulties, anxiety, depression, diabetes, angina, respiratory conditions, and arthritis (Van Cleemput, 2012). However, underreporting, and premature death make it impossible to understand the true incidence and impact (Parry et al, 2007).

Variations in lifestyle within the GTRC may create varying degrees of inequality within the population: The gypsy and traveller in the UK have maintained a more nomadic way of life and are more likely to live in caravan-based accommodation and sites compared to European Roma communities (Brown and Scullion, 2009). The UK gypsy and traveller environment and lifestyle is potentially more conducive to health problems, contributing to a greater disease burden (Smith and Ruston, 2013). Many factors present challenges such as a history of mistrust of authorities and literacy and language difficulties (Kelleher et al, 2011). The nomadic lifestyle can make accessing social-care services and engagement with medical and health authorities more challenging, making health promotion interventions more difficult meaning that problems are more likely to unnoticed, unreported, and untreated (Smith and Ruston, 2013).

#### Socioeconomic deprivation

The burden of poor health is markedly greater for those in the most socioeconomically (SE) deprived groups: Those in the poorest social class have been found to have a 60% higher prevalence of long-term, non-communicable diseases, such as cardiovascular disease (CVD), diabetes, and chronic obstructive pulmonary disease than those in the richest social class (Department of Health, 2012). Those in SE deprived groups have 30% more severity of disease (Department of Health, 2012), as well as poorer disease outcomes (Sharrocks et al, 2014). Greater SE deprivation increases the likelihood of multiple conditions (Taskforce on Multiple Conditions, 2021), to the extent that over a quarter of people suffering highest deprivation have more than 4 long-term conditions and they develop multiple long-term conditions around 10 years earlier than those in the least deprived population (Stafford et al, 2018).

In terms of specific diseases there is a considerably higher incidence of cancer in SE deprived groups (Donnelly and Gavin, 2011), to the extent that more than 30,000 extra cases of cancer in the UK each year are attributable to SE deprivation, and survival is worse for the most deprived groups (CRUK, 2022).

Those in the most deprived areas are four times more likely to die of CVD compared to the most affluent areas (PHE, 2019), CVD is also more common in those with severe mental illness or those from south Asian or African Caribbean populations (PHE, 2019). Compounding facets in this complex situation are that Asian people are the most likely out of all ethnic groups to live in the most deprived areas, followed by Black people (Gov.uk, 2020) and mental illness has a higher incidence in areas of SE deprivation (Remes et a, 2019), a situation which is being exacerbated by the cost of living difficulties (ONS, 2022a).

A 10-year study shows a worsening relationship between deprivation and poor health: Life expectancy is diminishing, with the greatest slow-down occurring in the most deprived populations (Marmot et al, 2020). Data on avoidable deaths; classified as those which could have been avoided through timely and effective healthcare interventions such as public health and primary prevention, show that in 2020 avoidable deaths accounted for 40.1% of all male deaths in the most deprived areas in contrast to 17.8% in the least deprived areas; for female deaths, it was 26.7% and 11.9%, respectively, and was found to have worsened over the past year (ONS, 2022b).

#### Homeless people

Homeless people experience extreme inequity across a wide range of physical and mental-health conditions, with a mortality rate 12 times higher than the general population. Factors which largely contribute to this are accidents, substance abuse/overdoses (Aldridge et al, 2018). Research data to understand the health issues and prevalence of many diseases among homeless people in the UK are lacking (Bowen et al, 2019, Al-Shakachi et al, 2020). Perplexingly, the prevalence of some diseases such as CVD, stroke, diabetes, and cancer, appear lower, however this is likely to be a consequence of underreporting or incorrect coding at times of healthcare interactions (Bowen et al, 2019). Furthermore, the author suggests that, comparable to the phenomenon seen in the GTRC, premature death is likely to distort the data on disease prevalence.

Socioeconomic and lifestyle factors such as smoking, alcohol and diet contribute to greatly increased non-communicable disease risk in this population (Al-Shakachi et al, 2020). A SR by Al-Shakarchi et al (2020) found that homeless people have a three times higher risk of CVD and increased CVD mortality compared to the housed population. Electronic health record data from over 40,000 homeless people over a period of 19 years revealed a greater incidence of CVD and quantified that comorbidities and risk-factors were more prevalent in homeless people (Nanjo et al, 2020). This observational data reinforces the need to expand research in this area to inform pathways for treatment and prevention.

### What are the effects and implications of underrepresentation in clinical research

#### Gypsy, Traveller, Roma

Those in the GTRC suffer a greater disease burden and health disparity compared to all other ethnic minority groups and those living in other similar socioeconomic circumstances (Condon et al, 2021a). However, lack of research data creates an inaccurate picture of the incidence and progression of diseases. For example, the incidence of cancer and disease progression among the GTRC is not well known or understood due to scarcity of research data, deeming it an ‘invisible’ health concern (Condon et al, 2021b). Parry et al (2007) found lower rates of cancer reported amongst the GTRC as evaluated against age-sex matched comparators. However, given the socioeconomic and lifestyle risk factors (Cook et al, 2013, ONS, 2014) along with reduced accessing of preventative health services (Peters et al, 2009) it is suggested that this deceptively low incidence is a result of late presentation, lower life expectancy or reduced survival rates (Van Cleemput, 2019). This indicates that a lack of reliable data means that screening and treatment cannot be provided, targeted, or promoted appropriately.

#### Socioeconomic deprivation

The phenomenon of under-representation of the SE deprived population has been documented (Bonevski et al, 2013). A complicating factor is that the term SE deprivation encompasses myriad other underrepresented groups. Hence, there may be some degree of inclusion of other population groups not specifically intended for inclusion in this SR, who are, as a result, considered as SE deprived, such as migrant populations and some ethnic minorities. For this SR to focus upon the included study population the author has ensured to only consider papers with a prevalent and primary concern of SE deprivation.

Underrepresentation in the SE deprived population means that medical data inaccurately reflects the population, safety of health innovations are poorly understood, and diseases which present the highest burden within this population are poorly understood in terms of occurrence and progression (Bonevski et al, 2013).

A population level multi-variate analysis to explore participation in cancer research trials found that those in the most deprived quintile were least likely to participate in a clinical research trial (Donnelly et al, 2017). Cancer Research UK (2020) state that as well as higher incidence of cancer and poorer outcomes there are disparities in cancer treatments between highest and lowest deprivation levels, despite similar patient and disease characteristics. The reasons for this are unclear, hence they implore that the focus of research is directed towards those known to be at greater risk and least likely to survive.

A quantitative analysis of a CVD risk trial in the SE deprived population found that those at higher risk were least likely to attend a research-led programme of screening (Lang et al, 2015). The implication of this is lack of knowledge and evidence regarding correlation between causative factors. It is estimated that reducing the gulf in uptake of screening and prevention in the highest risk groups could eliminate almost 70% of the excess deaths in the SE deprived population (Harkins et al, 2010). This would help the immediate population and would provide data to understand the associations more deeply between SE deprivation and cardiovascular risk and how best to tackle the health inequalities which are known to exist (Lang et al, 2015).

#### Homeless people

There is a paucity of data on the homeless population in relation to specific physical diseases and progression. This is partly because of lower levels of routine screening and healthcare pathways, therefore the main source of data on healthcare encounters is from Accident and Emergency attendances (Bowen et al, 2019). The collection of data using electronic health records proved that there are risks and disease burdens in this population but due to the lack of health research evidence there is little known to inform health pathways and interventions. This means that screening programmes, and preventative measure are poorly informed and specific guidelines are lacking for homeless healthcare services (Al-Shakarchi et al, 2020).

### Why certain barriers and facilitators may exist

#### Gypsy, Traveller, Roma

Combined factors concerning lifestyle, insular culture, and history of marginalization and prejudice make it challenging to access, recruit, and retain participants from the GTRC (Van Cleemput, 2018). Research participation is typically viewed with suspicion due to experiencing oppression (Smith, 2021). To compound these factors further the GTRC tend to have a fatalistic approach to health (Van Cleemput et al, 2007).

The logistical difficulties around identifying and engaging eligible participants with nomadic lifestyles are understandable (Van Cleemput, 2018), nonetheless even with those living on authorised sites engagement can still be challenging (Brown and Scullion, 2010). Therefore, when forming relationships between researcher and community it is key that the community can participate in research in an empowered way, rather than them being the subject of research. It has been reported that this can cause the community to feel they are the subject of a consultation, sometimes repeatedly, as a ‘well-meaning’ way of addressing research inequity which leads to disillusionment (Brown and Scullion (2010).

Social networks and social capital are powerful factors within the GTRC, with a strong culture of reciprocity and sharing of health information (Smith and Ruston, 2013). Rather than viewing this as a barrier by viewing the community as hard-to-reach, if is suggested that this can be harnessed and capitalised upon as a key to facilitating engagement within the GTRC (Brown and Scullion, 2010).

#### Socioeconomic deprivation

Historically, low recruitment in this group has been reportedly due to issues of culture and mistrust (Bonevski et al 2014). With the label of being ‘hard-to-reach’ or ‘hard-to-access’ inferring that the population make themselves elusive to the researcher (Sydor, 2013). However, it is increasingly recognised that the label of hard-to-reach tends to be imposed upon individuals or groups due to socioeconomic determinants of health, discrimination, or exclusion rather than this being a choice made by the participant (Belt et al, 2022). Sharrocks et al (2014) report several fundamental issues within the fabric of the research process which present as barriers for this population, such as eligibility criteria for clinical trials, referrer wariness and hesitancy, and the language and readability of research materials.

Bonevksi et al (2014) report several studies which cite that practical and financial aspects can present barriers to recruitment due to work, childcare and transport challenges. Whilst financial incentives and reimbursements can facilitate greater participant satisfaction and retention (Bonevski et al, 2014), the bioethical perspective must ensure that incentives are not coercive and should not be used as a prime strategy to overcome engagement and recruitment barriers (Resnik, 2023). Consequently, the practical need for reimbursement should balance with other prominent themes of community engagement, building relationships with participants and showing courtesy, care, and compassion (Bonevski et al, 2014), and by employing socially and culturally proactive and innovative approaches (Sharrocks et al, 2014).

#### Homeless people

Dominant barriers for recruitment and retention are the practical issue of locating individuals living on the street (Bonevski et al, 2014), and the lack of safe and appropriate places to recruit and conduct research activities (Ojo-Fati et al, 2017). Mental health difficulties and substance misuse are frequent reasons for exclusion to research trials (Aldridge et al, 2018), therefore Ojo-Fati et al (2017) advocate flexibility within trials schedules, and flexibility around eligibility and inclusion.

Mistrust of authority and apprehension of seemingly intrusive questions can pose barriers in this group particularly when regarding illegal behaviour (Bonevski et al, 2014). However, as within their SR and meta-analysis Aldridge et al (2018) conducted an engagement workshop showed which showed 100% willingness within the homeless population for the collection and use of hostel data to support a health inclusion initiative.

### What has been done so far and initiatives for the future

Historically, a systematic approach aimed at understanding and overcoming underrepresentation in research trial populations has been lacking (Witham et al, 2020b). Through the lessons learnt from research during the covid-19 pandemic intentions have become polarised with the objective of commitment and collaborations between NHS England, National Institute for Clinical Excellence (NICE), National Institute for Health Research (NIHR) to deliver the shared aims of increasing public participation in research, and to engage more diverse research participants across health and social care (Gov.uk, 2021).

Through the NIHR key guidelines and national initiatives have developed and evolved with the aim of achieving this objective. The NIHR INCLUDE guidance was conceived in 2017 as one of a suite of significant projects aiming to address inequities in representation and promote inclusivity in health and care research (NIHR, 2020). The project was completed in 2021 and has been absorbed by the overarching NIHR Underserved Communities’ project. The vision and direction for delivery of this project is presented in the Best Research for Best Health operational guidance (NIHR, 2021).

Subsequently, the NIHR Equality, Diversity and Inclusion strategy (NIHR 2022a) was formatted. Following on from this, a pilot, entitled Research Ready Communities (2022b), was conducted in three local underserved areas, using community champions to partner with local voluntary and community sector organisations. This pilot was completed late 2022 and proved so successful in engaging under-served communities and improving inclusion and representation that it was suggested to be adopted nationally and used as a broad framework to be adapted locally and flexibly depending on need. The author envisages that this SR will help to guide what the need is, what approach and resources may be required for it to be delivered most appropriately, and how the role of community champions can be developed and expanded upon.

### Methodology

This study uses SR methodology, an approach which provides the basis for evidence-based healthcare (Moola et al, 2015) by extracting, appraising, and synthesising all data relevant to the research question (Munn et al, 2014). This SR will explore the barriers and facilitators affecting participation in health research trials amongst socio-economically disadvantaged populations in the UK and Ireland.

This SR of etiology, also known as SR of association (Moola et al, 2015), will include quantitative and qualitative research over the past 10 years in the UK and Ireland to provide a narrative synthesis (NS) (Stern et al, 2021).

This SR was conducted according to PRISMA guidelines (Page et al 2021). The use of PRISMA is not to assess methodological quality of reviews (Sarkis-Onofre et al, 2021), but is to ensure a clear, comprehensive, and accurate report of why the review was done, what was done, and what was found (Page et al, 2021).

### Inclusion Criteria

The study will use a PEOS framework.

Population: Populations of SE deprivation, gypsy, traveller and Roma communities, and homeless people, within the UK and Ireland.

Exposure: Those from the defined population groups who have participated in health research and/or where data have been obtained or provided on factors regarding participation in health research trials.

Outcomes: Reported factors affecting participation in health research, including barriers & facilitators.

Study design: The SR will include primary research papers of qualitative and quantitative study design, conducted between 2013-2023.

### Systematic literature review

As is apparent from the literature review there is evidence to demonstrate underrepresentation of certain groups in health research. A traditional literature review presents a general overview of a topic, demonstrates the development of knowledge about a topic, identifies where evidence may be lacking or inconclusive and justifies why a subject or issue warrants further study (Aromataris and Pearson, 2014). However, they can tend to be subjective and rely on the authors experience, and do not provide such extensive evidence and unbiased synthesis as SR (Davies, 2019).

The intention of SR is to follow a rigorous and structured approach to obtain the most valid evidence to develop reliable guidelines and inform decisions (Aromataris and Pearson, 2014). A suggested limitation of SR is that traditionally they focus upon exploring the effectiveness of medical interventions (Munn et al, 2018). Pearson et al (2015) contend that effectiveness alone is not always the best way to address a research question and advocate a more pluralistic approach. A further criticism of SR is that it lacks epistemological reflection and presents a lower degree of scientific originality and decreased methodological rigor in comparison with primary research (De los Santos et al, 2022).

These criticisms of SR have led to the formation and development of an array of frameworks and approaches to address and answer the question in the most appropriate way possible (Munn et al, 2018). This SR endeavours to address potential weaknesses perceived within the SR methodology by conducting an SR of etiology to provide a NS of the evidence to establish a critical overview of the relevant research findings over the last 10 years. By using the chosen approach and methodology this supports the notion that narrative SR can be valid and methodical provided it adheres to rigorous and transparent standards (Gough et al., 2019).

These chosen methodological approaches and design allow an interpretivist approach, to counteract the criticism that SR is ostensibly weighted towards evidence-based medicine and clinical decision making (Gough et a, 2019), traditionally with a mainly positivist approach (De los Santos et a, 2022).

A SR of etiology was chosen to determine associations between exposures, risk factors and outcomes within the defined population, which enhances understanding of the relationship of health-related outcomes by exploring associations between variables (Moola et al, 2015).

Typically, in SR the PICO (population, interventions, comparators, and outcomes) format is used for study eligibility criteria, however this aligns less well with SR of etiology (Moola et al, 2015). Therefore, the PEOS format was considered preferable for exploring associations between exposures/factors and outcomes (Munn et al, 2018).

Inclusion of qualitative and quantitative research was chosen to achieve a balance of data based on positivism and interpretivism (De los Santos et al, 2022). By using a mixed approach, the aim is to focus on the research question in a real-life context via multi-level perspectives (Pearson et al, 2015).

NS presents the findings as a summary of knowledge, described as a whole, rather than describing each study in turn. This was chosen to allow an exploration of associations, differences, and relationships between the findings of the studies and to observe for any patterns within the findings, which may otherwise less obvious (Ryan, 2013).

## Methods

### Search strategy

The search was conducted using electronic databases only. The databases used were EBSCO host, MEDLINE, CINAHL Ultimate, CINAHL, and Psychology and Behavioural Sciences Collection. These were searched systematically for articles published between 2013 and 2023 using search terms, limiters, and expanders, presented in Appendix 1.

Two pilot searches were run using individual criteria within the search terms, this gave a small yield of results. Therefore, the search strategy was modified to utilise the PEOS mnemonic. This allowed the author to firstly focus on all of populations which were intended to be included. The exposure was defined as being exposed to a health research trial, crucially, with an element which examined reasons participation or non-participation in. The outcome was defined as whether participation or recruitment occurred or not and whether the article referred to barriers or facilitators concerning this.

### Application of inclusion and exclusion criteria

The selection of papers was guided by the PEOS format. This checklist further defines and expands upon the criteria for inclusion and exclusion.

#### Population

- The study was completely or partly related to any or all the defined population groups, within the UK or Ireland.

Exposure:

- The study related to clinical, medical or health trials, where one or more of the defined population groups participated in research and/or where data were obtained or provided relating to factors around participation in health research trials.

#### Outcome

- The study reported evidence, outcome or discussion regarding barriers or facilitators to recruitment or health research participation.

#### Study Design

- The study was qualitative or quantitative primary research.
- The study was conducted between 2013-2023 (within the last 10 years).
- The study was available in English language.

### The PEOS selection template

Titles and abstracts were first screened for inclusion/exclusion, then the full paper was screened. A template was used (Appendix 3) to apply the above criteria to each individual paper. This was adapted from a PEOS template suggested by Bettany-Saltikov and McSherry (2016). This allows the next level of the selection process where papers will be recorded with more detail, before fully appraising the methodological quality of the papers.

### Data extraction

Data extraction is essential to synthesize and combine data from various studies and is fundamental to the way in which the SR can extend beyond a being a narrative summary and ensures that data extracted from individual research studies are relevant to the SR question (Munn et al, 2104). The data extraction form was designed and piloted to emphasize the characteristics of the studies and illustrate how the relevant data were extracted in a standardised format.

### Validity assessment

Appraising study quality and validity is intended to assess methodological quality of the primary papers selected for SR and to consider to what extent they are likely to represent the truth (Bettany-Saltikov and McSherry, 2016). The purpose of assessing the selected studies is not usually to exclude studies at this stage but rather to be able to appraise the impact which the quality of the studies have upon the reliability of the results and therefore on the conclusions drawn (Centre for Reviews and Dissemination, 2009). Typically, quantitative methodology appraisal is most concerned with validity and reliability, whereas quality studies are concerned with authenticity and trustworthiness (Bettany-Saltikov and McSherry, 2016). Reviewing a combination of both quantitative and qualitative papers involves different factors and is therefore important to use a process which can attend to both aspects (Aromataris and Pearson, 2014, Caldwell et al, 2011).

For these reasons, the author chose to use and adapt the framework and questions developed by Caldwell et al (2011) for the critical appraisal.

A flow chart to illustrate the questions for evaluation of validity for both qualitative and quantitative is in Appendix 4.

The author made individual scoring tables separately for qualitative and for quantitative papers. A score of 0, 1 or 2 was allocated to each criterion, meaning that a maximum score of 36 could be given across the 18 questions. For quality appraisal scoring it is difficult to find a consensus on how recent the literature reviews within the selected papers should be (Snyder, 2019). Bettany-Saltikov and McSherry (2016) indicate that studies which include literature reviews with superior balance and relevance to the research topic should be marked more highly. Therefore, it was decided to award higher scores for papers with literature review references largely within the last 10 years, with exceptions for key seminal references.

### Data analysis

To aid the data analysis process the author used a colour-coding system for each study paper using one colour to highlight facilitators and another to highlight barriers, this information was then transcribed into the table, which recorded the study characteristics and presented the identified barriers and facilitators regarding engagement, recruitment, and retention within research trials for the population group.

### Data synthesis

The data synthesis uses a NS approach. The purpose of this is to conduct a textual approach that produces an analysis of the associations within and between studies and an overall assessment of the strength of the evidence (Popay et a, 2007). The nature of NS can potentially lean towards a subjective process compared to methods such as meta-analysis, hence, the approach used should be rigorous and transparent to limit the potential for bias (Popay et al, 2007). The Cochrane guidance is that SR should ideally be conducted with a second reviewer, as this was not possible the Cochrane guidance on data synthesis was followed to minimise bias as much as possible (Ryan et al, 2013).

Once the main themes were identified through data extraction and NS, a basic data matrix table was used to show presence, absence, and overlap of themes across the three population groups. These were then summarised narratively by group, with commonalities and differences within and between groups highlighted.

## Results

The initial searches generated a yield of 263 titles and abstracts. After the search engine removed duplicates, this reduced to 194. The results were exported to an Excel spreadsheet. An initial sift manually removed papers which were listed twice due to variation in bibliographic formatting, but which were the same paper in the same journal. This provided a list of 181 articles to which the inclusion and exclusion criteria listed were applied. This is illustrated in the PRISMA flowchart in fig 1. This excluded 168 papers based on title and abstract, a list of excluded papers is in appendix 2. This left 13 remaining papers to be fully read. All 13 remaining papers which were read fully met all inclusion criteria, therefore there were no further exclusions.

**Fig. 1.**
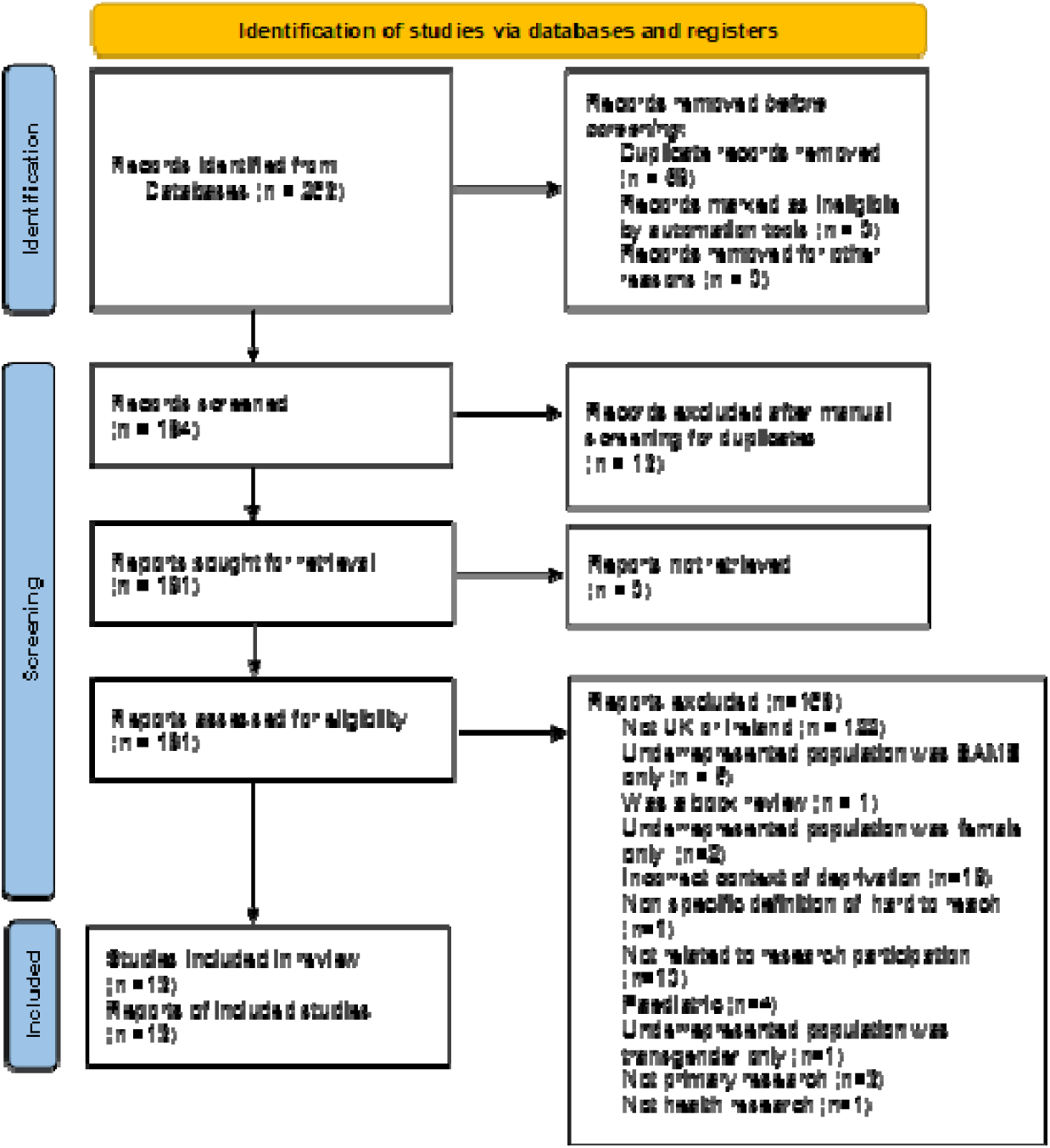
PRISMA flow chart.

Table 1 presents a summary of the PEOS features of the 13 remaining selected papers.

**Table 1:** Study characteristics using PEOS criteria.

| Table 1: Study characteristics using PEOS criteria. |  |  |  |  |
| --- | --- | --- | --- | --- |
| Author, year | Population | Exposure | Outcome | Study design/ methodology |
| Astbury et al, 2020. | SE deprivation. N=272 | Uptake to clinical trial of a diet replacement weight loss programme, via GP invite letter for those in areas of SE deprivation | Assess which participant characteristic factors may affect uptake and clinical trial enrolment in the community | Quantitative, Cochran-Mantel-Haenszel analysis. Randomized 1:1 – intervention programme: usual care |
| Biswell et al, 2021 | Homeless. N=64 | Experiences of clinical research nurses and homeless patients when recruiting and participating in the 'homeless hospital research study'. | Explore experiences of homeless people in a research study concerned with discharging homeless people from hospital and the challenges experienced by clinical research nurses. | Qualitative case-study discussion paper based on the authors' experiences. |
| Condon et al, 2019 | GTR N=50, across 4 sites | Exploration and discussion of challenges and opportunities to engage gypsy, traveller, and Roma communities in a multicentre qualitative research project | Explore factors around public involvement and community engagement to provide insights into conducting effective research in this community | Multi-centre, qualitative interview |
| Condon et al, 2022 | GTR N=6 | Reflections on conducting peer interviews as part of a participatory research project exploring experiences of cancer within the gypsy/traveller/Roma community | Explore the role and the challenges of PPI when conducting cross cultural research in the community related to a sensitive health topic in marginalised groups | Qualitative, peer interviews as a part of a participatory research project |
| Dewhurst et al, 2022 | SE deprivation. N=100, across 3 sites | Opportunities for palliative care inpatients to take part in research trials and how the relationship between prognosis and socioeconomic status may affect their wish for involvement in research trials | Explore levels of interest and preferences of palliative care hospital inpatients concerning research trial participation and any relationship between study design demographics and prognosis | Quantitative, multicentre cohort study questionnaire |
| Islam et al, 2021 | SE deprivation. N=48, across 5 researchers and 12 community organisations. | Public and community involvement and engagement in translational research planning to improve co-design of trials in communities of high BAME and lower socioeconomic status. | Explore and address issues which are experienced around underrepresentation of BAME and low socioeconomic status groups. To level the power relationship between communities, researchers, and research infrastructure | Community sandpit. Predominantly qualitative researcher interviews and community organization representative focus group data. Combined with quantitative questionnaire. |
| O'Reilly-de Brún et al, 2016 | SE deprivation. N=51 | Participatory learning and action (PLA) to explore issues of healthcare research exclusion in migrants known to experience low-income and economic difficulties | Explore PLA as a normative part of healthcare research in the community to incorporate views and needs of the defined population into the research agenda | Qualitative, PLA |
| Quaife et al, 2019 | SE deprivation. N=2012 | Comparison of effects of different strategies in uptake of lung screening trial based on SE status | Assess how targeted research trial invitations in the community impacts under representation of people of low socioeconomic status | Quantitative, Two-arm blinded RCT |
| Rogers et al, 2019 | SE deprivation. N=1050, across 58 sites | Use of different clinical trial recruitment approaches in clinical trials, based on demographics including socioeconomic deprivation | Assess the impact and effect of different invitation strategies based on SE deprivation levels | Quantitative randomised trial |
| Smyth et al, 2020 | GTR N=3 | Use of scripted performances and discussions in health research | Assess the impact of creative holistic approaches in improving engagement in health research in marginalised populations in the community | Qualitative, pilot of a modified version of Boal's simultaneous dramaturgy |
| Tully et al, 2021 | SE deprivation. N=86 | Recruitment strategies with underrepresented, marginalized populations to build rapport and engagement with health research | Assess the impact of multi-pronged approaches in enhancing engagement with health research in the community | Qualitative, focus groups |
| Vickneson et al, 2021 | SE deprivation. N=11,059 | Factors influencing participation and commitment to a research trial based upon demographics including SE deprivation levels | Measure how medical factors and SE factors may influence recruitment, retention, and compliance in a remote clinical trial | Quantitative, large remote randomised trial |
| Walsh et al, 2022 | GTR and homeless. N=30 | Participatory approaches aimed at improving engagement and representation of marginalised populations in health research and the development and implementation process | Provide critical reflections on the experiences of marginalised community populations and how they are represented in health research | Qualitative, participatory voice-led methodology (interview, focus-group and forums) |

These remaining papers were evaluated for quality and scored using the 18 questions from the Caldwell et al (2011) quality evaluation framework, appendix 4. Scores of 0, 1, or 2 were awarded, meaning a maximum possible score of 36. The eight qualitative papers achieved scores between 19 and 34. The five quantitative papers achieved scores between 32 and 35.

The combined scores for quantitative and qualitative papers are in Appendix 5.

To extract the data each paper was examined for characteristics and to identify themes of barriers and facilitators for research participation. The tabulated data extraction is in appendix 6. A tabulated summary of the themes of potential barriers and facilitators for each population group are presented in Appendix 7. From these tables the NS was developed.

### Description of included studies

Thirteen studies were identified, of which 8 were qualitative and 5 quantitative. Twelve studies were concerned with one population group, one study was concerned with more than one population group. Eight studies were SE deprivation (Astbury et al, 2020, Dewhurst et al, 2022, Islam et al, 2021, O’Reilly-du Brun et al, 2016, Quaife et al, 2019, Rogers et al, 2019, Tully et al, 2021, Vickenson et al, 2021). Three studies were gypsy/traveller/Roma (Condon et al, 2019, Condon et al 2022, Smyth et al, 2020). One study was homeless Biswell et al, 2021). One study was both homeless and gypsy/traveller (Walsh et al, 2022).

The populations, setting/exposure, outcome or aim of the study and the study design are detailed in table 1. Eleven studies took place in a community setting, one was initiated in a hospital setting and then community-based follow-up (Biswell et al, 2021), one was solely hospital based (Dewhurst et al, 2022). All papers across the populations achieved evenly spread evaluation scores of between 27-36, except for one qualitative paper for the GTRC, with 3 participants (Smyth et al, 2020), which scored only 19.

### Gypsy/traveller/Roma community

There were four studies about the gypsy/traveller/Roma community; all were qualitative studies, consisting of one multi centre interview study (Condon et al, 2019), one peer-researcher interview study (Condon et al, 2022), one pilot of a modified version of Boal’s simultaneous dramaturgy (Smyth et al, 2020) and one participatory voice-led study using interview, focus-group and forums (Walsh et al, 2022).

### Barriers for the gypsy/traveller/Roma community

#### Cultural, sensitivity and identity

One paper reported that distinctions of subgroups within the whole group risked causing offence and posed the problem for the gatekeeper that they were not seen as impartial (Condon et al, 2019). The Showpeople subgroup had some reluctance to participate as they didn’t consider themselves aligned with the other subgroups (Condon et al, 2019), showing the problem of outsider classifications with the groups. Shared ethnicity with the peer-researcher, whilst largely considered a facilitator, was also a barrier when talking one-to-one about culturally and emotionally sensitive topics (Condon et al, 2022)

#### Location and practical issues

There were common themes of location and setting: In one study (Smyth et al, 2020) the initial setting for the event was a university theatre, no one attended at the first attempt, thought to be because it was not a place visited by the community. No roadside gypsy/travellers were recruited, potentially due to practical issues of the location of the interviews (participants homes or community venues) and in cases where there was a less well-established relationship with local gatekeepers it made it difficult to secure a suitable location (Condon et al, 2019). Walsh et al (2022) reported practical difficulties beyond the location challenges in so much as structural uncertainties and disrupted lifestyle hindered engagement and commitment as the population tend to focus on one day at a time. Walsh et al (2022) also reported that lack of access to technology and unfamiliarity with online platforms was a hindrance.

#### Gatekeepers

Where gatekeepers were involved, they predominantly assisted engagement, therefore the nature of the role is described subsequently in greater detail in the context of facilitators. However, Condon et al (2019) reported that where there were concerns that the gatekeeper was not impartial or where gatekeeper relationships were less well-established this presented a barrier. Additionally, barriers were created due to local authority cuts resulting in reduced availability of gatekeeper in some sites (Condon et al, 2019)

#### Language and literacy

One study reported that being non-English speaking was an initial barrier to recruitment, Smyth et al, (2020) were unable to obtain a Romani translator, however some assistance was given by one of the participants who was able to translate. Literacy issues were a challenge especially in the older age groups, which made text-based aspects difficult, with participants stating that being given a lot of forms made them feel stigmatised and embarrassed (Walsh et al, 2022). Smyth et al (2020) noted that written consent was a problem due to literacy problems and that audio recording of verbal consent could have been a preferable option.

#### Mistrust

Despite the interviews being conducted by peer-researchers the use of an audio recorder caused the participants feelings of concern about confidentiality and created a degree of mistrust and reluctance to speak freely due to the realisation that this would be shared with others (Condon et al, 2022),

#### Health issues

One study reported issues of poor attendance, leaving early due to anxiety, withdrawal due to poor physical health, although possibly intensified by the fact that this study was specifically concerned with older adults (Walsh et al, 2022).

### Facilitators for the gypsy/traveller/Roma community

#### Relationships and trust

The benefits of fostering and utilising relationships of trust between gatekeepers, community representatives/leaders, peer-researchers and participants were a common theme within two of the four papers (Condon et al 2019, Walsh et al, 2022). The gatekeepers were from various backgrounds; charities, service providers and representative groups (Walsh et al, 2022), and familiar trusted members of the community (Condon et al, 2019). Gatekeeper sensitivity and awareness of social and political allegiances was beneficial in successfully placing subgroups within the overall study population (Condon et al, 2019). Walsh et al (2022) reported that gatekeeper involvement in recruitment protocol development and in facilitating recruitment was key to the success of their study.

Although shared ethnicity was a potential barrier particularly with sensitive topics, overall, it enhanced feelings of advocacy and assurance of confidentiality (Condon et al, 2022), created a non-threatening, respectful approach and the participants valued shared recognition of injustices faced by the study group (Condon et al, 2019). Engagement and recruitment were enhanced through a face-to-face cooperative approach with open continuous dialogue and providing chances for participants to ask extra questions, including being able to explore researcher credentials (Smyth et al, 2020). Social and personal benefits and drivers to engagement were that participants valued the opportunity to meet with others, share experiences, exchange stories and situations (Walsh et al, 2022), and valued access to additional information regarding their health (Smyth et al, 2020).

#### Location and practical issues

Familiarity and comfort of the venue enhanced engagement (Smyth et al, 2020), and flexibility to take research to homes or community venues and provide drop-in options lead to greater inclusion and engagement (Condon et al, 2019). Condon et al (2019) also identified that flexibility to be able to accommodate and include other family members, even if not required for the sample, maximised engagement. Participants valued the opportunity to suggest and involve others to take part and this was found to enhance engagement and participation (Smyth et al, 2020). Practical aspects such as provision of a voucher (Condon et al, 2019) and provision of food and drink (Smyth et al, 2020) were facilitating factors. Walsh et al (2022) reported that gatekeepers supported practical aspects of recruitment and retention by providing technical assistance with emails and video links for the interviews.

#### Language and literacy

The ability to overcome language barriers was beneficial, not just in practical terms of sharing the same language (Condon et al, 2022), but all the studies noted the wider cultural benefits in terms of reciprocal dialogue and understanding of nuances within language and discourse (Condon et al, 2019, Smyth et al, 2020, Condon et al, 2022, Walsh et al, 2022). Some peer-researchers found that more highly educated participants were more willing to talk at length, whereas lower levels of education resulted in briefer more clear-cut responses and realised the need to accept this and value whatever information they got, not to push too much and risk losing engagement, illustrating the benefit of understanding variations within the study population (Condon et al, 2022).

### SE deprived group

Eight studies were about the SE deprived group, five were quantitative studies, consisting of four randomised controlled trials (Astbury et al, 2020, Quaife et al,2019, Rogers et al, 2019, Vickenson et al, 2021) and one multi-centre cohort study (Dewhurst et al, 2022). One was a predominantly qualitative study of researcher interviews and community organization representative focus-group data, and researcher observations with the addition of a quantitative and qualitative questionnaire (Islam et al, 2021), one was a qualitative focus groups study (Tully et al, 2021), one was a qualitative participatory learning and action study (O’Reilly-de Brun et al, 2016).

### Barriers for SE deprived groups

#### Relationships and trust

Only one paper reported issues of trust as a barrier (Islam et al, 2021), stating that participants felt a stigma around the term and the label of being ‘hard-to-reach’, when they actually considered it was hard to trust the researcher. From the researchers’ own practice observations, it was reported that lack of knowledge and understanding of the research process and the role of diversity and inclusion within research results in reduced participation in research. It was specifically reported by participants that not knowing what was expected of them being in a research trial made some feel uncomfortable and reluctant (Islam, et al, 2021).

Community leaders and gatekeepers were largely a facilitating factor, which will be reported subsequently. However, one study reported that barriers were created where there were challenges in building initial rapport and maintaining buy-in with some gatekeepers (Tully et al, 2021). Tully et al (2021) also reported the concept that the presence of the gatekeeper potentially created a ‘double edged sword’ in that they could feel protective over their community group and wanted to protect them from being perceived as inadequate.

#### Financial and resource issues

Three quantitative studies measuring deprivation index reported a direct correlation with the highest levels of SE deprivation resulting in lower levels of recruitment and commitment to research trials (Quaife et al, 2019, Rogers et al, 2019, Vickenson et al, 2021). Vickenson et al (2021), specifically reported that not possessing the necessary equipment requirement for participating in the trial, in this case a validated home blood pressure monitor, led to lower compliance and lower long-term commitment in the most deprived participants. Actual or perceived financial factors, such as loss of income during participation or actual or perceived costs incurred were reported as a barrier from a researcher observation on one study (Islam et al, 2021). Tully et al (2021) found that despite offering travel reimbursement no participants took this up, they reflected that those who were unable to pay upfront would not have joined the study, they would therefore consider arranging transport for a future study.

Three studies did not identify any barriers to participation in the SE deprived population (Astbury et al, 2020, Dewhurst et al, 2022, O’Reilly-de Brun et al, 2016)

### Facilitators for SE deprived groups

#### Health related issues

Two of the quantitative study results opposed the common trend of reduced uptake within the most SE deprived groups (Astbury et al, 2020, Dewhurst et al, 2022). Astbury et al (2020) explored participant characteristics and relationship to uptake and retention in a weight loss study and reported higher uptake in those in the highest and intermediate deprived tertiles and those with the highest BMI, which they suggest could be due to the study being more appealing to those for whom the health issues were most pertinent. Similarly, the study by Dewhurst et al (2022) also countered the trend of reduced uptake in SE deprived groups. They compared deprivation levels to see if this affected willingness to take part in palliative care research and reported that desire to participate was not affected by SE deprivation status.

#### Relationships and trust

All three qualitative studies reported and described positive aspects of trusted relationships. O’Reilly-de Brun et al (2016) reported on using service-user peer-researchers from within the population group to conduct the research by providing a link between university researchers and the community. As this study encompassed SE deprived economic migrants, shared language was a key facilitator, which, combined with cultural concordance enabled complex participatory learning and action research and achieved high levels of recruitment and retention. The participants reported feeling motivated and empowered, they valued being able to have their own say and share their knowledge and opinions and felt honoured to participate, and it reduced their feelings of isolation. Similarly, Islam et al (2020) reported that positive partnerships were formed between community groups and researchers by using trusted community figures and leaders. Participants reported that this created a ‘safe-space’ and a ‘level playing field’ by balancing power dynamics between researchers and communities. (Islam et al, 2020). The study by Tully et al (2021) was largely enabled by using community organisations, reporting that 91% of participants were recruited via this route, whereas only 9% were via social media. Achieving buy-in and building rapport with gatekeepers involved in community organisations was key to successful recruitment. This was found to be more productive and effective when giving the gatekeepers as much information as possible, and speaking by phone rather than email proved more beneficial (Tully et al, 2021)

#### Screening and recruitment issues

Simplifying the approaching and recruiting process proved a common theme as a facilitator across two quite divergent studies. A large scale RCT of >2000 subjects found that targeted, staggered approaches gave higher levels of uptake (Quaife et al, 2019). Similarly, a qualitative focus group study found that reducing steps within screening and approaching and allowing participants to self-screen improved uptake and recruitment (Tully et al, 2021).

#### Location issues

One qualitative study reported benefits of the familiarity of the location (Tully et al, 2021), specifically flexibility with timings and arranging a research event around a pre-existing community event.

#### Financial and reimbursement

There were no reports of specific financial reimbursement, so it was not reported to be a facilitator. However, in one quantitative study it was suggested that provision of total diet replacement products as part of the trial, which would usually cost around £700, seemed more appealing to those with limited finances (Astbury et al, 2020).

#### Language and literacy

For SE deprived participants in a qualitative study involving economic migrant shared language with the peer-researcher created rapport and engagement (O’Reilly-de Brun et al, 2016). Low burden materials were the most effective in engaging the most deprived individuals, suggested to correlate with lower levels of literacy (Quaife et al, 2019).

Two studies did not identify any facilitators in the SE deprived population (Rogers et al, 2019, Vickenson et al, 2021).

### Homeless people

There were two qualitative studies about homeless people. One was a participatory voice-led study using interviews, focus-groups, and forums to explore how this marginalized population can be integrated into research on health and ageing (Walsh et al, 2022). The other was a case study discussion based on the authors’ experiences of a homeless health and social care research project, which considered barriers and facilitators from both researcher and participant perspective (Biswell et al, 2021).

### Barriers for homeless people

#### Relationships and trust

Both papers reported that suboptimal researcher/participant relationships affected trust, engagement, recruitment, and retention. Despite using gatekeepers in a highly collaborative and representative approach one study unfortunately failed to recruit people currently living on the street, although there were participants who had had recent experience of this (Walsh et al, 2022). Biswell et al (2021) specifically reported that having to asking uncomfortable questions relating to arrest or prison felt uncomfortable for the recruiter and created suspicion and feelings of mistrust from the participant perspective and were unsure how these questions related to their health.

#### Practical recruitment and location challenges

Biswell et al (2021) reported that researchers having to work outside their usual familiar setting required more time to build relationships with service-users. They also reported challenges in identifying research recruits to approach due to that fact that homeless status is not routinely recorded in hospital data. Once participants were recruited there were difficulties in locating them for follow-up, and follow-ups were time-consuming.

#### Health and lifestyle

Both papers reported similar overlapping challenges regarding health and lifestyle. The focus on surviving one day to the next (Walsh et al, 2022) and the reality of frequently changeable sleep sites (Biswell et al, 2021) create barriers to engagement and commitment. As well as poor attendance there were also instances of leaving early due to anxiety and/or poor physical health (Walsh et al, 2022). Periods of emotional and/or mental health crisis and/or intoxication made answering questions difficult or impossible, furthermore, substance misuse meant certain times of day were not feasible or suitable to approach participants (Biswell et al, 2021).

#### Literacy

One paper reported challenges with literacy and cognition, which may have been compounded by being an older demographic (Walsh et al, 2022). The participants reported that being given numerous forms which they didn’t understand caused embarrassment. Walsh et al (2022) also reported that participants would either not have access to or be unfamiliar with online platforms or technology and lacked support networks to assist them with this.

### Facilitators for homeless people

#### Practical and location

Biswell et al (2021) reported that from the researcher perspective it was an advantage to have some clinical research background as it aided their familiarity with the workplace environment and systems, and an understanding of medical/health conditions and circumstances meant that the approach to the participant could be done more smoothly. Walsh et al (2022) reported that gatekeepers supported practical aspects of recruitment and retention by providing technical assistance with emails and video-links for interviews to be conducted.

#### Financial and reimbursement

Due to the research nurses’ knowledge and familiarity with the hospital environment they were able to address unmet needs of homeless patients on the ward by knowing how to access resources within hospital to be able to provide clothing, toiletries and to wash their dirty clothes (Biswell et al, 2021). This was an action over and above the design of the research study, however it resulted in considerable gratitude and engagement from the participants. Within the study reported by Biswell et al, (2021) there was provision of a £10 shopping voucher as a thank you for participation, the benefits were not explicitly reported, but as many participants came to hospital without any money, it was expected that this would be valuable to them.

#### Relationships and trust

Biswell et al (2021) reported that the status of the research nurse was important in fostering relationships with community services which assisted in locating participants, hence was beneficial for the researcher. Regarding participant benefits both papers reported on the value of relationships and trust, whether in the role of a gatekeeper or a research nurse. The role of the gatekeeper in the community setting was key in building trust and relationships and understanding the nuances of the population group (Walsh et al, 2022). In the hospital setting knowledgeable research staff meant participants not having to repeat themselves which helped to overcome suspicion, put participants at ease and created a foundation to establish a relationship (Biswell et al, 2021). Furthermore, the compassion and kindness from research nurse encouraged participation and resulted in gratitude. Both studies reported feelings of motivation and reward from the participants which encouraged engagement and participation. The participant felt the research topic was important and they valued the chance, particularly at follow-ups, to talk about their experiences (Biswell et al, 2021). Within the focus groups the participants valued the opportunity to be able to meet up to talk about their experiences, exchange stories and situations (Walsh et al, 2022)

#### Language and literacy

The papers reported that equality of dialogue (Walsh et al, 2022) and use of accessible language (Biswell et al, 2021) enabled participants to discuss health within the research and enhanced interest and commitment from participants. Where literacy and cognition difficulties created challenges for text-based research activities, images and visual aids were used and were reported to enhance engagement (Walsh et al, 2022).

### Overlap of barriers and facilitators

Table 2. illustrates the extent of overlap of reported barriers and facilitators across the populations.

**Table 2.** Matrix of overlap of barriers and facilitators.

| **Table 2. Matrix of overlap of barriers and facilitators** |  |  |  |
| --- | --- | --- | --- |
| **Barriers** | **Gypsy/traveller/Roma** | **SE deprivation** | **Homeless** |
| Difficulties with education/literacy | X |  | X |
| Difficulties with Language/dialogue | X |  |  |
| Inadequate/inappropriate location | X |  | X |
| Reduced access to/familiarity with technology | X |  | X |
| Lack of understanding of research process and expectations |  | X |  |
| Mistrust, or challenges with gatekeeper/researcher relationships | X | X | X |
| Challenges around cultural sensitivity/sensitive topics | X |  |  |
| Financial/material challenges |  | X |  |
| **Facilitators** | **Gypsy/traveller/Roma** | **SE deprivation** | **Homeless** |
| Positive relationships and/or rapport and trust in gatekeeper/research staff | X | X | X |
| Financial/material incentives | X | X | X |
| Social benefits/sharing stories | X | X | X |
| Feelings of empowerment or sense of reward/value | X | X | X |
| Connectivity through language/dialogue | X | X | X |
| Allowances for variations or challenges with education/literacy | X | X | X |
| Benefits to health | X | X | X |
| Flexibility and adaptability in recruitment participation | X | X | X |

As illustrated in the matrix there is a complete overlap of facilitators across all populations. Whereas fewer barriers were reported, notably from the SE deprived group and therefore fewer overlaps.

## Discussion

A range of barriers and facilitators across the groups were identified from this SR.

### Common themes in barriers between groups

Eight key themes emerged across the groups: ‘difficulties with education/literacy’, ‘difficulties with language/dialogue’, ‘inadequate/inappropriate location’, ‘reduced access to/familiarity with technology’, ‘lack of understanding of research process and expectations’, ‘mistrust, or challenges with gatekeeper/researcher relationships’, ‘challenges around cultural sensitivity/sensitive topics’, ‘financial/material challenges’.

Only three barriers in total were identified for the SE deprived group. This could be because the SE deprived group had a greater proportion of quantitative papers; five out of the eight. Whereas the studies of the GTRC group and homeless group were all qualitative papers, which enabled a richer identification and description of the themes.

Two barriers of ‘lack of understanding of research process and expectations’, and ‘financial/material challenges’ were unique to the SE deprived group, whereas ‘cultural sensitivity/sensitive topics’ and ‘difficulties with language/dialogue’ were unique to the GTRC group.

Four of the barriers identified applied to more than one group. GTRC and homeless were the populations which shared the largest number of common barriers of ‘difficulties with education/literacy’, ‘inadequate/inappropriate location’ and ‘reduced access to/familiarity with technology’. These barriers lend themselves to practical solutions, which, as was reflected in the initial literature review should be addressed by ensuring flexibility in conducting research (Ojo-Fati, 2017) and by employing socially and culturally proactive and innovative approaches (Sharrocks et al, 2014).

The single common barrier shared across all three groups was that of ‘mistrust, or challenges with gatekeeper/researcher relationships’. This theme had commonalities across all groups in terms of a barrier being created where there was absent or inadequate trust or rapport between the researcher/gatekeeper and the participant. The initial literature review referenced mistrust as a longstanding major barrier for underserved groups (Bonevski et al, 2014). The fact that this continues ten years later demonstrates the pervasive nature of this issue and reflects the need for on-going, innovative, and progressive work to continue to address this across all groups. Parallels can be drawn to a recent study reported by Ekezie et al (2021), which demonstrated a prevalence of similar themes across different underserved groups terms of mistrust, misinformation, and apprehension regarding covid-19 vaccine research. They advocate that where issues may appear the same, when they are arising from different population groups the approach should be targeted to the individual group (Ekezie et al, 2021).

The GTRC and SE deprived papers both reported that having peer-researchers from within the population and/or shared ethnicity proved to be both barrier and facilitator. Within the SE deprived group where there were gatekeepers/peer-researchers it was reported that they could tend to feel protective over their peers and want to protect them from potential stigmatisation by being a research participant. In the GTRC group where the gatekeeper/researcher and participant shared the same ethnicity it was reported that the participant found it difficult to discuss emotionally sensitive health or societal issues. Within the initial literature review it was reported by Brown and Scullion et al (2010) that the strength of culture, reciprocity, and social networks in the GTRC should be harnessed to enable engagement, rather than be viewed as a barrier. However, the challenges identified in this SR around discussing delicate topics with peer-researchers of shared ethnicity illustrates the power of culture within this group and the challenges which that presents.

### Common themes in facilitators between groups

Eight key themes emerged across the three groups, each of these were common to all three groups. Many of these facilitators correlate in some way to relationships, emotions, and connections: ‘positive relationships and/or rapport and trust in gatekeeper/research staff, ‘social benefits/sharing stories’, ‘feelings of empowerment or sense of reward/value’, ‘connectivity through language/dialogue’. Whereas some have a more practical aspect to them: ‘allowances for variations or challenges with education/literacy’, ‘benefits to health’, ‘flexibility and adaptability in recruitment participation’, and ‘financial/material incentives’.

Although there were commonalities across all groups, there were variances both across different groups, and within homogenous groups. In the GTRC it was found that distinctions existed within the groups in terms of language and dialogue: The UK/Ireland gypsy/travellers have certain types of dialogue where it proved beneficial for the peer-researchers to have shared knowledge and insight to break down cultural barriers, whereas some of the Roma gypsies had a different language which required use of an interpreter. There was also some overlap of these factors across heterogenous groups: Within one of the SE deprived studies, some of the populations were economically challenged migrants and therefore, akin to the Roma population, had a different language, hence required an interpreter. The homeless groups valued equality of dialogue and use of accessible language comparable to some degree with the UK/Ireland gypsy and traveller groups.

The most prominent facilitator across all the studies which was described and discussed in the most detail and depth was that of relationships. It could be suggested that the permeation of positive relationships creates an effect of perpetuating facilitating factors, and without the presence of positive relationships and connections the practical solutions may be less manifest as facilitating factors. This is reflective of outcomes from a previous SR by Bonevski et al (2014), as they reinforced that tackling practical barriers must be wholly balanced with engagement and building relationships with respect, care, and compassion.

### Strengths and limitations of the studies

The qualitative papers for GTRC and homeless had greater focus on roles of peer-researchers, participatory approaches and community engagement which produced greater breadth and depth of information.

The fact that factors relating to gatekeeper relationships and peer-researchers were both facilitator and barrier, and was the only common barrier across all groups, indicates the value of participatory approaches in partnership between community and researchers as a way of ensuring that these relationships are as optimal as possible.

The identification of barriers and facilitators for the SE groups was weaker than for homeless and GTRC groups. This was despite there being a larger number of papers for the SE deprived group. Two of the quantitative papers for the SE deprived group failed to identify any facilitators. Two quantitative papers and one qualitative paper for the SE deprived group failed to identify any barriers.

Overall, the identification of facilitators proved much stronger compared to identification of barriers. This raises the concern that the absence of barriers may be because the most marginalised people who experience the most marked barriers have failed to be reached or engaged. A contributary factor in the lack of identifying barriers may be that the quantitative papers had more focus toward demographic indicators to research uptake rather than exploring the causes for lack of uptake. The fact that there were fewer barriers identified from the quantitative trials cannot wholly be taken to mean that these factors do not exist, as they were not actively explored, and it is more difficult to evidence an absence of a factor in this case.

### Quality and validity of the evidence base

The aim of the mixed methods SR was that the research question could be explored in in a real-life context via multi-level perspectives. A varied range of methodologies were included, particularly with the qualitative papers. Evaluating and synthesising the literature in a balanced manner was challenging to some extent due to the methodological variations in the papers, despite using an evaluation framework intended for both quantitative and qualitative studies. For example, the qualitative studies presented variations ranging from simultaneous dramaturgy with three participants through to multi-centre studies with between 50->80 participants. The quantitative studies had variations ranging from cohort studies of 100 subjects through to large scale RCTs with >11,000 participants. The SE deprived studies were heavily weighted towards quantitative studies which failed to qualify what the barriers were and lacked detail behind the facilitators which were identified. Within the quantitative SE deprived studies there was use of deprivation indices and areas of deprivation, which is less good at ascertaining individual deprivation status of participants. The Caldwell framework was chosen due to its suitability for mixed methods, however further validity evaluation tools were not explored. Further consideration of alternative frameworks would ensure that thorough consideration has been given to identifying the optimum framework for the SR.

### Methods strengths and weaknesses

Despite using methods guided by Cochrane and an established quality evaluation framework, only being able to use one reviewer within the SR means that some errors or bias may have occurred, which may have been avoided with the use of a second reviewer.

It was decided not to impose a lower limit for the quality evaluation score, which led to the inclusion of what could be considered lower-level evidence. However, whilst it may be difficult to draw irrefutable conclusions from the qualitative data obtained from the lowest scoring study, it provided valuable real-life insights, whereas the higher scoring large scale RCTs lacked certain aspects of evidence. Whilst not diminishing the value of any of the studies included in the SR, the variation and breadth of data could be improved by obtaining a wider array of studies and methodologies across the groups. This could have been achieved by accessing studies from outside of UK and Ireland but limiting the search to high income countries, to ensure it is as generalisable as possible to the UK.

### Implications for current policy and future approaches

All facilitators were common across all groups, albeit for different reasons. This continued focus should be on upon relationships and engagement according to the varying reasons to maximise the other recognised facilitators to full effect. This is reflective of previous findings in relation to community engagement in public health (Harden et al, 2015) where it was reported that the context of barriers and facilitators reflect pre-existing factors within communities. Furthermore, they reported that suboptimal relationships can create cynicism, cause feelings of or threat low motivation to engage and, simultaneously, engaging organisations are less likely to see the worth of community engagement. Where there are good relations, or efforts made to tackle longstanding poor relations, engaging organisations have supportive attitudes towards community engagement throughout the organisation and from the outset.

The lack of identified barriers suggests that more work should be done to learn what these may be for specific groups. The studies which reported facilitators had achieved what they had intended and demonstrated facilitators by the very nature of successfully engaging the populations. Whereas in the studies which failed to identify barriers, the prime aim had not been to discover what those barriers were, they focused more on illustrating a correlation. To explore and understand barriers there should be more work to locate and focus on those who have failed to be engaged. For example, one paper identified no people who were currently homeless and one failed to engage those with the worst levels of deprivation and in rural locations, therefore nothing is known about barriers and facilitators for these groups. This is reflective of previous reports on public health, that where engagement was low barriers or facilitators could not be discovered or understood (Harden et al, 2015).

Lessons from previous public health community engagement (PHE, 2021), combined with the findings of this SR, illustrate how the approaches of community champion roles and relationships can help to deliver the aims of the Research Ready Communities initiative (NIHR, 2022b). A previous review of community champion approaches demonstrates the value of reaching into target communities, including those which are often seldom heard, by using diverse and tailored approach (PHE, 2021), this approach would optimise the community research champion role to bridge the gap between researchers and community, in the most appropriate way for the community.

### Conclusion and Recommendations

It is valuable that some barriers were identified, however it may be prudent to further explore the overall absence of certain practical barriers particularly in the SE deprived group, such as financial issues and digital/technological barriers. It would seem likely that these exist as challenges in the populations. The fact that these were not reported may indicate lack of reach to the most deprived sections of this population. Therefore, more research needs to be done to fully explore the barriers in the most deprived groups.

In terms of future research engagement strategies, in the absence of being able to locate and engage these populations for research, it could prove valuable to draw parallels from retrospective studies of previously underserved groups, in both public health and health research, where community engagement has subsequently been effective. This could provide insights into what barriers had been experienced and how these had been overcome to inform how engagement with currently underserved groups can be developed.

The findings of this SR combined with policy and knowledge of public health and research initiatives indicate of the value of relationships, peer-researchers and community research champions in engaging with communities. It is hoped that by developing engagement approaches in a way which is tailored to different underserved groups this will address barriers and help to shape the future of research. There was a weighting towards community-based studies, however the author considers that it would be valuable for what has been learnt to be transferred from the communities and into the hospital-based clinical research setting. Presumably many barriers and facilitators which have been discussed within this SR will be present when a person becomes a hospitalised, possibly more so due to added fear, anxiety, and unfamiliarity. Therefore, it would be valuable for community research champions to explore, engage and prepare people for what may happen and how they may potentially feel if hospitalised and approached for research participation.

## Data Availability

All data produced in the present work are contained in the manuscript

## List of abbreviations

BAME: Black, Asian, and Minority Ethnic
BMI: Body Mass Index
CINAHL: Cumulative Index to Nursing and Allied Health Literature
CVD: Cardio-Vascular Disease
CRUK: Cancer research UK
EDI: Equality, Diversity, and Inclusion
EBSCO host: Elton B. Stephens Company
GTRC: Gypsy, Traveller, and Roma Community
MEDLINE: Medical Literature Analysis and Retrieval System Online
NICE: National Institute for Clinical Excellence
NIHR: National Institute for Health Research
NS: Narrative Synthesis
ONS: Office for National Statistics
PEO/PEOS: Population, Exposure, Outcome/Population, Exposure, Outcome, Study design
PHE: Public Health England
PICO: Population, Interventions, Comparators and Outcomes
PPIE: Patient ad Public Involvement and Engagement
PRISMA: Preferred Reporting Items for Systematic Reviews and Meta-Analyses
RCT: Randomised Control Trial
SE: Socio Economic
SR: Systematic Review

## Appendices

**Appendix 1:**
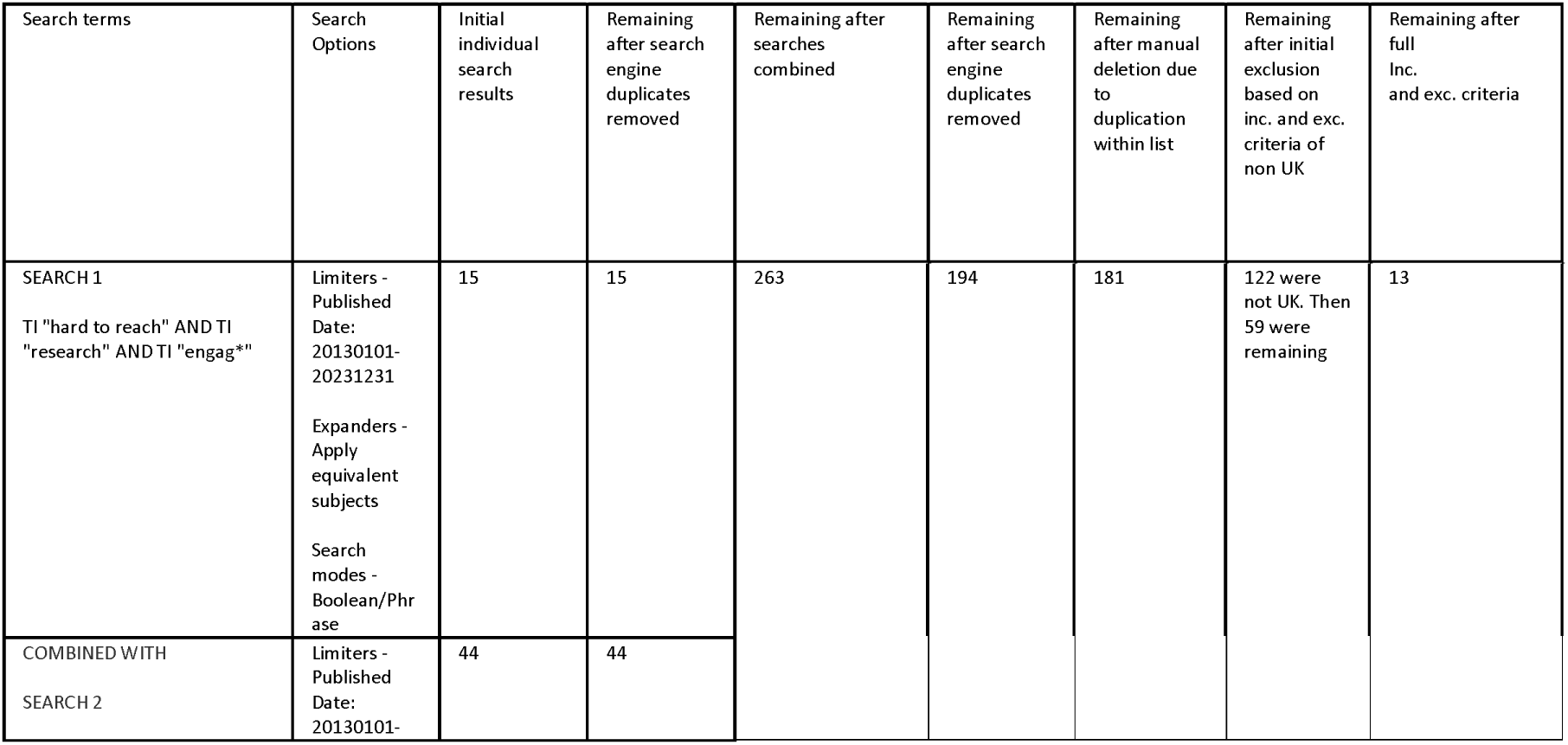
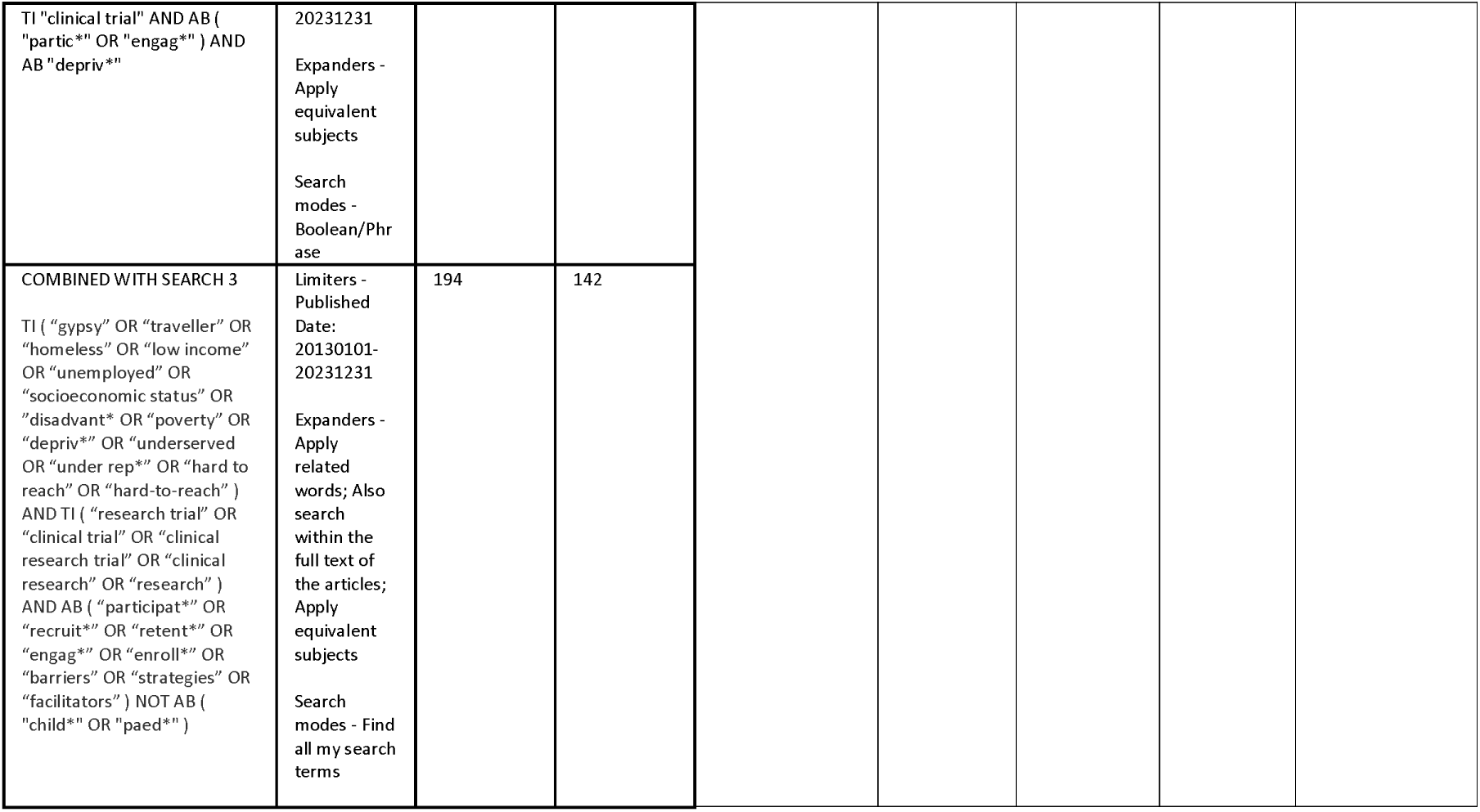
Search terms.

**Appendix 2:**
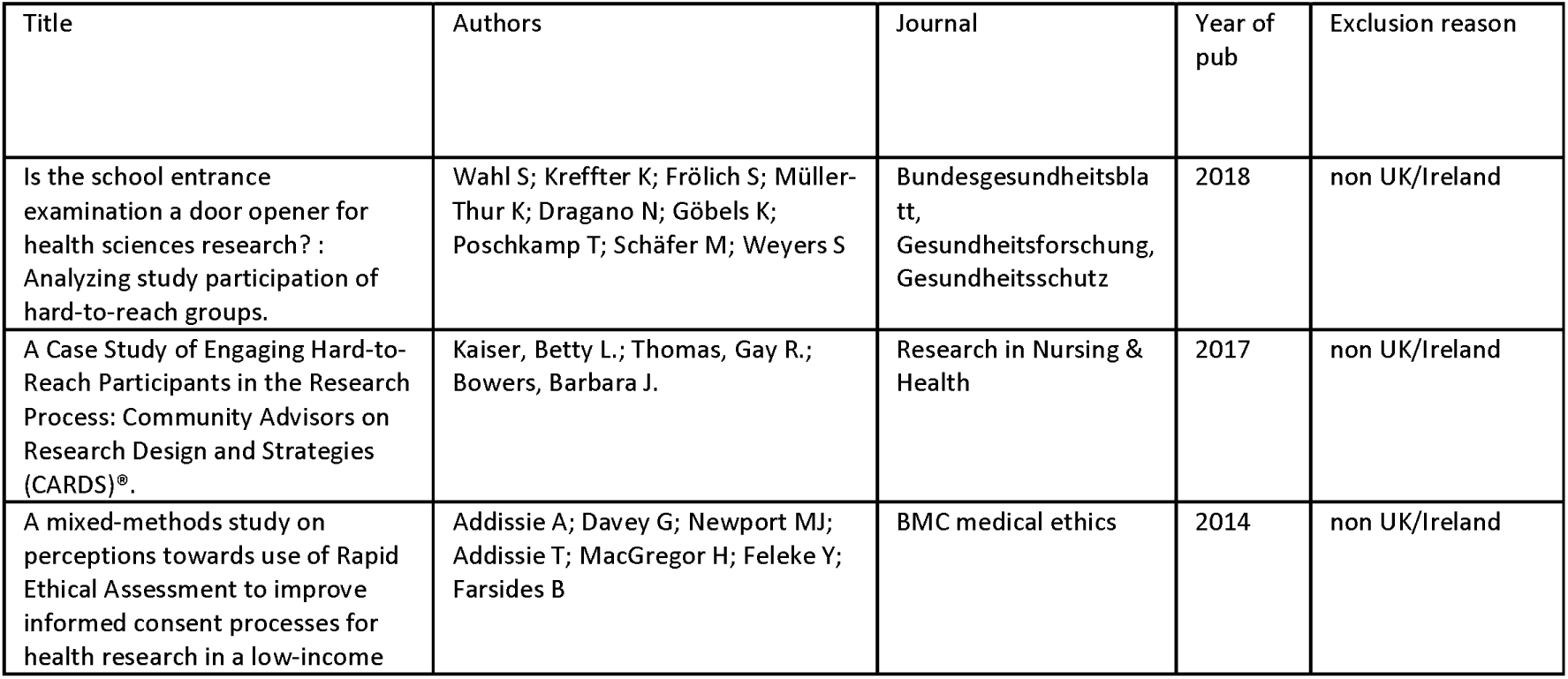
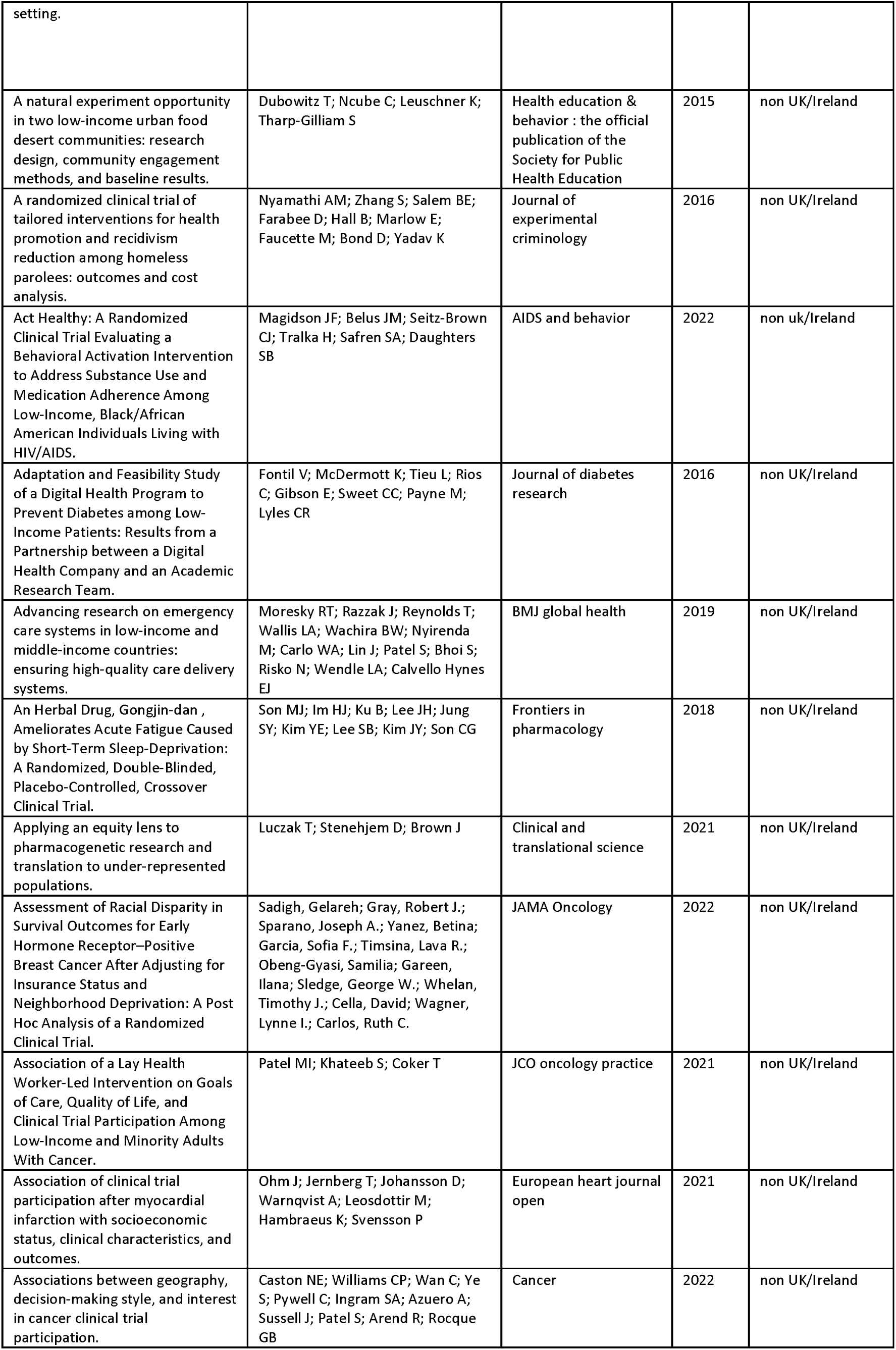
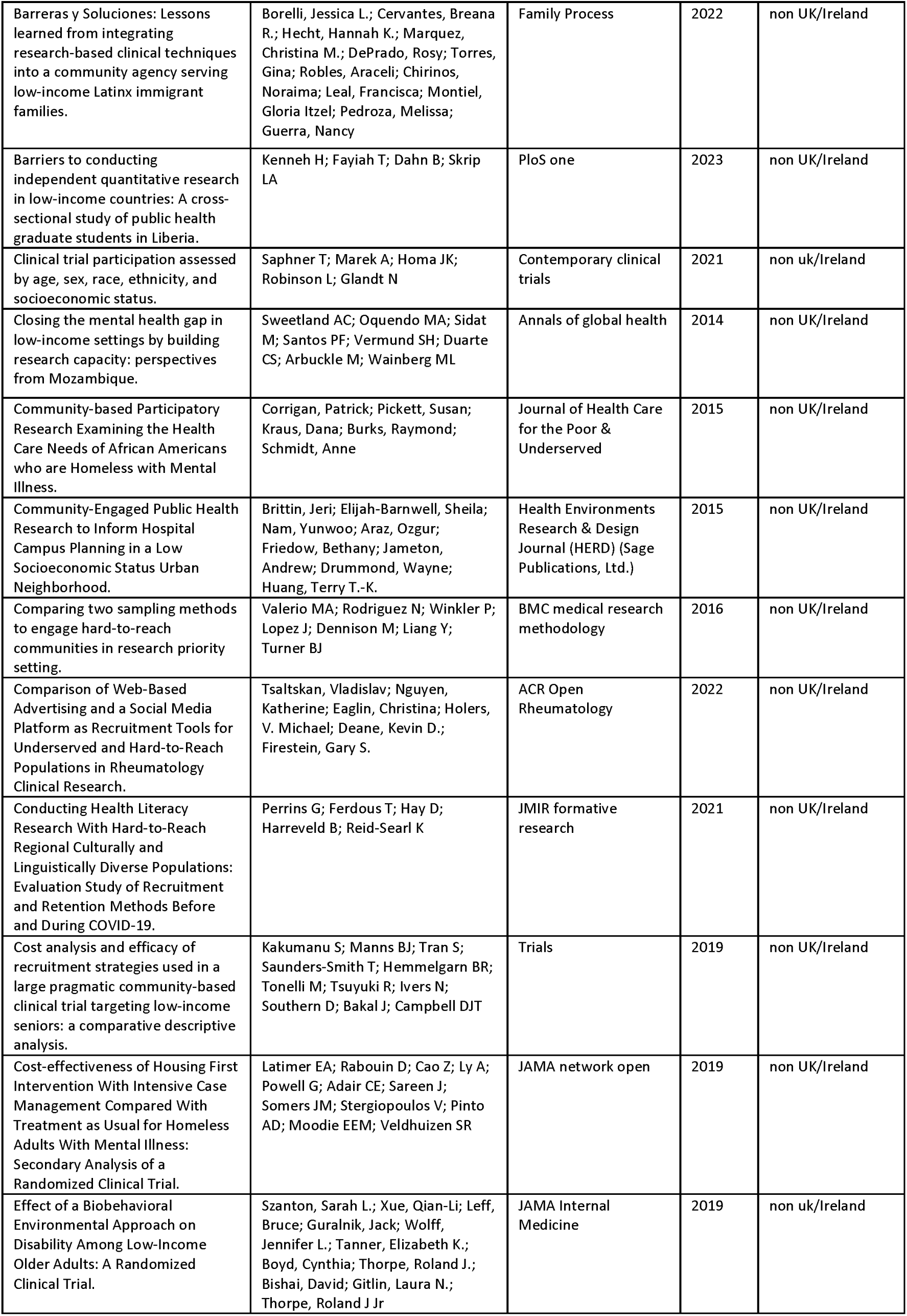
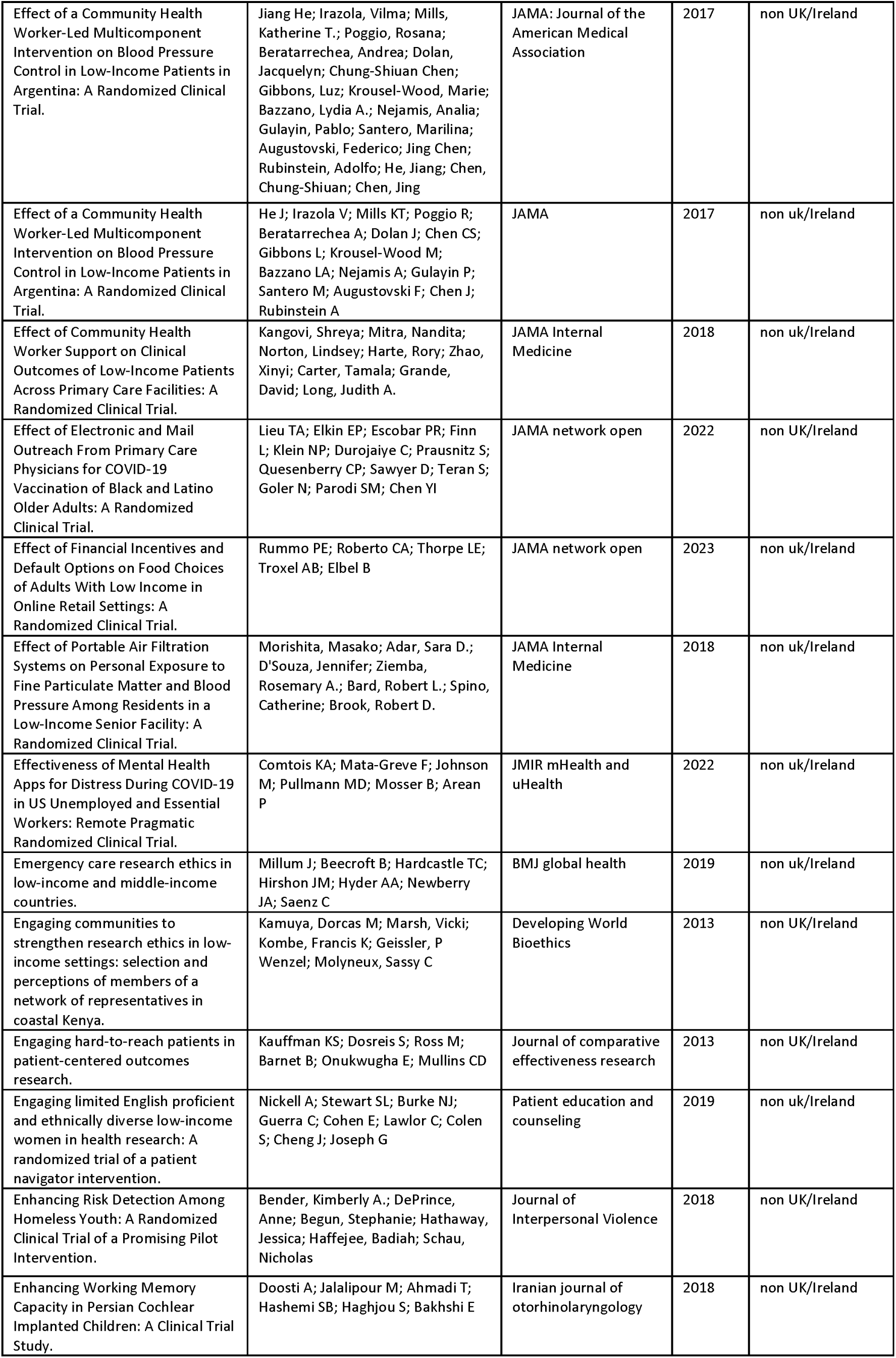
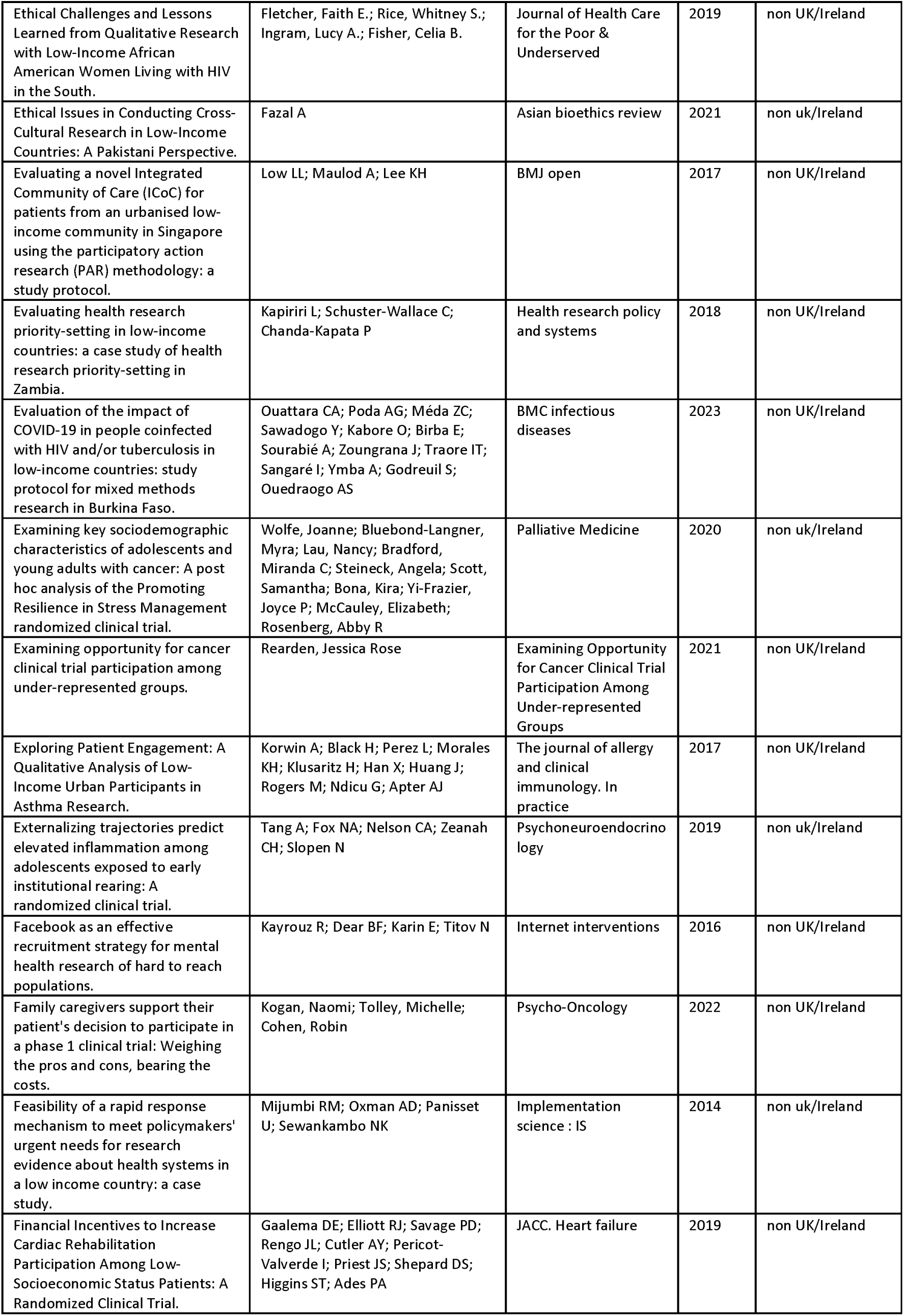
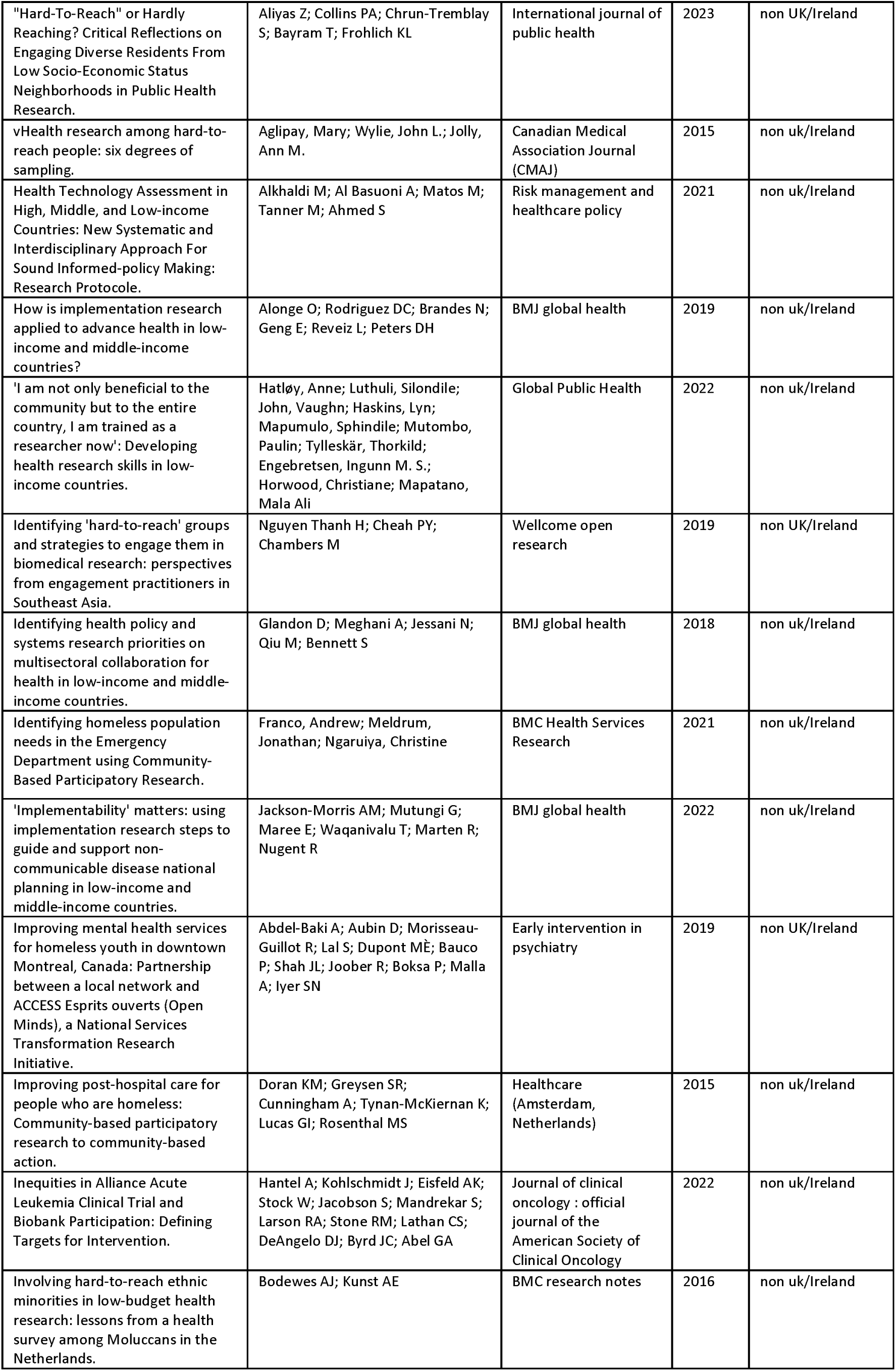
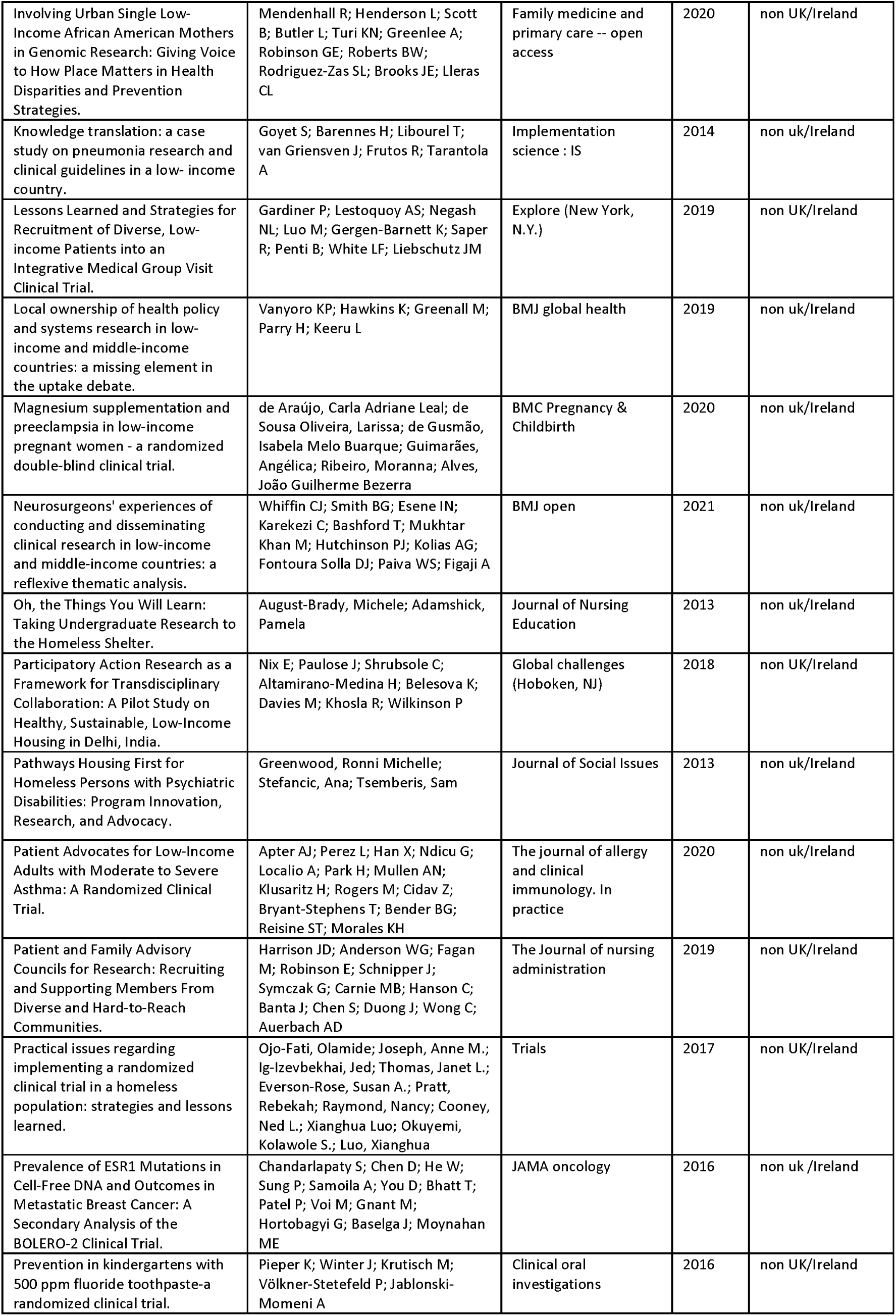
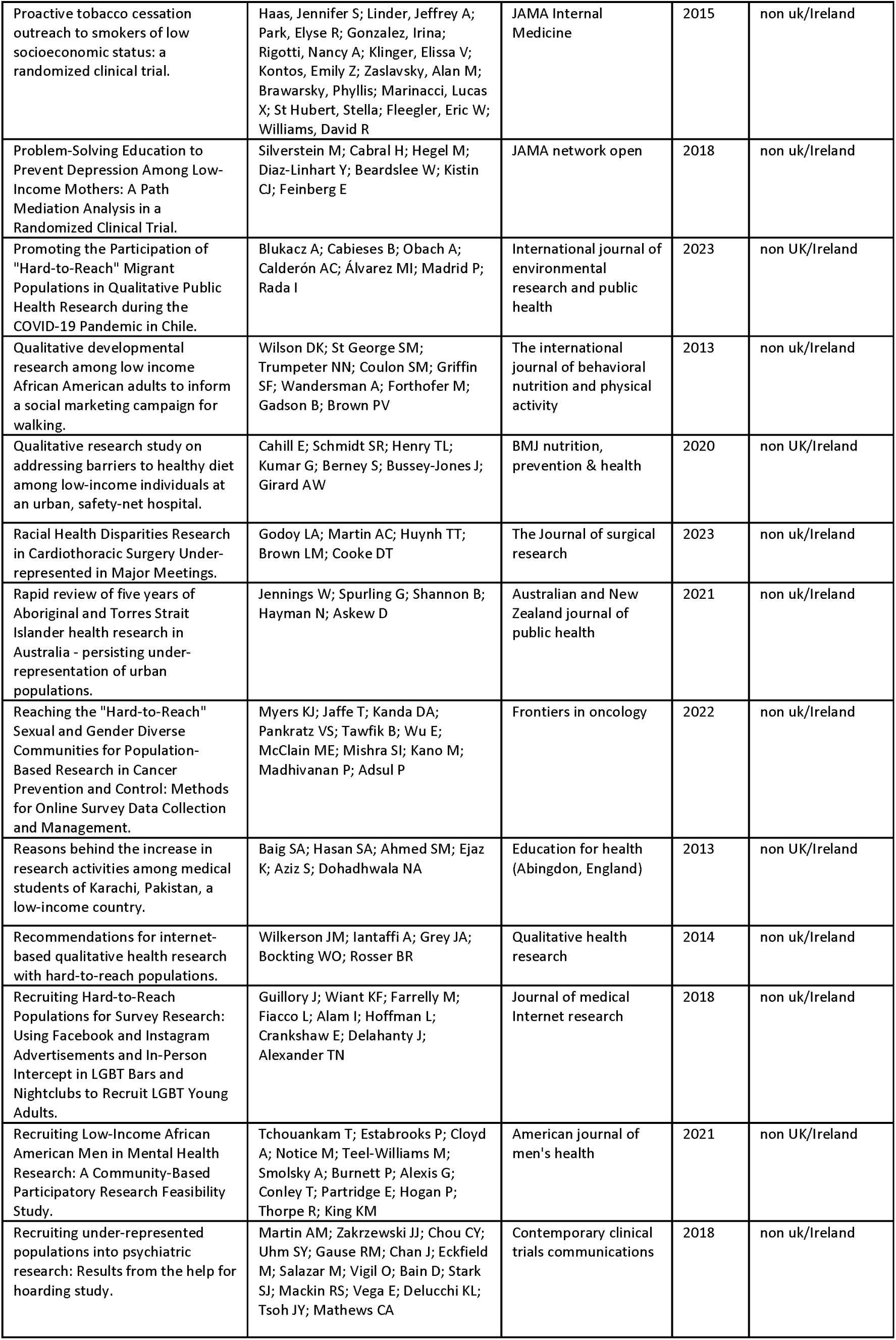
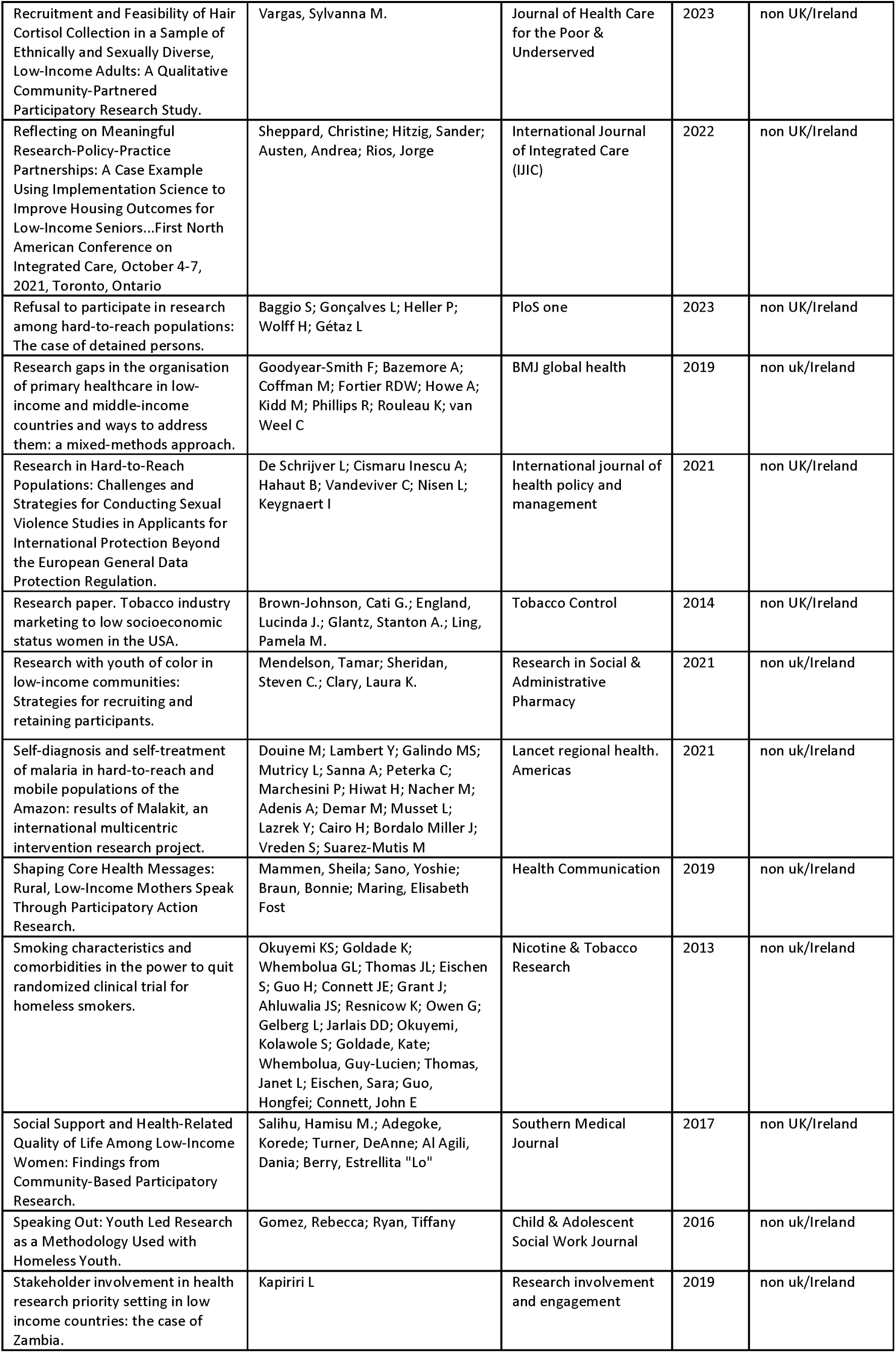
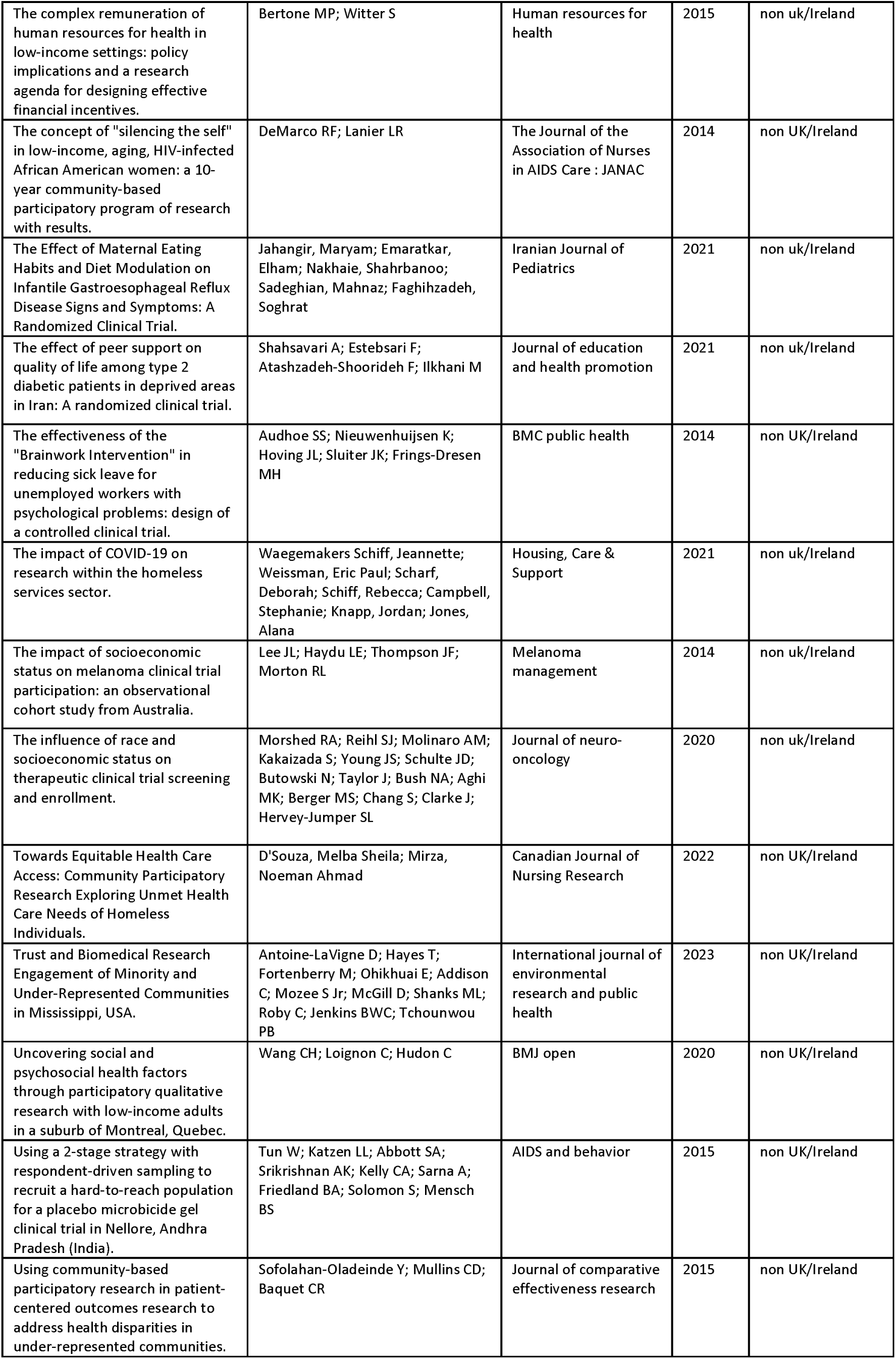
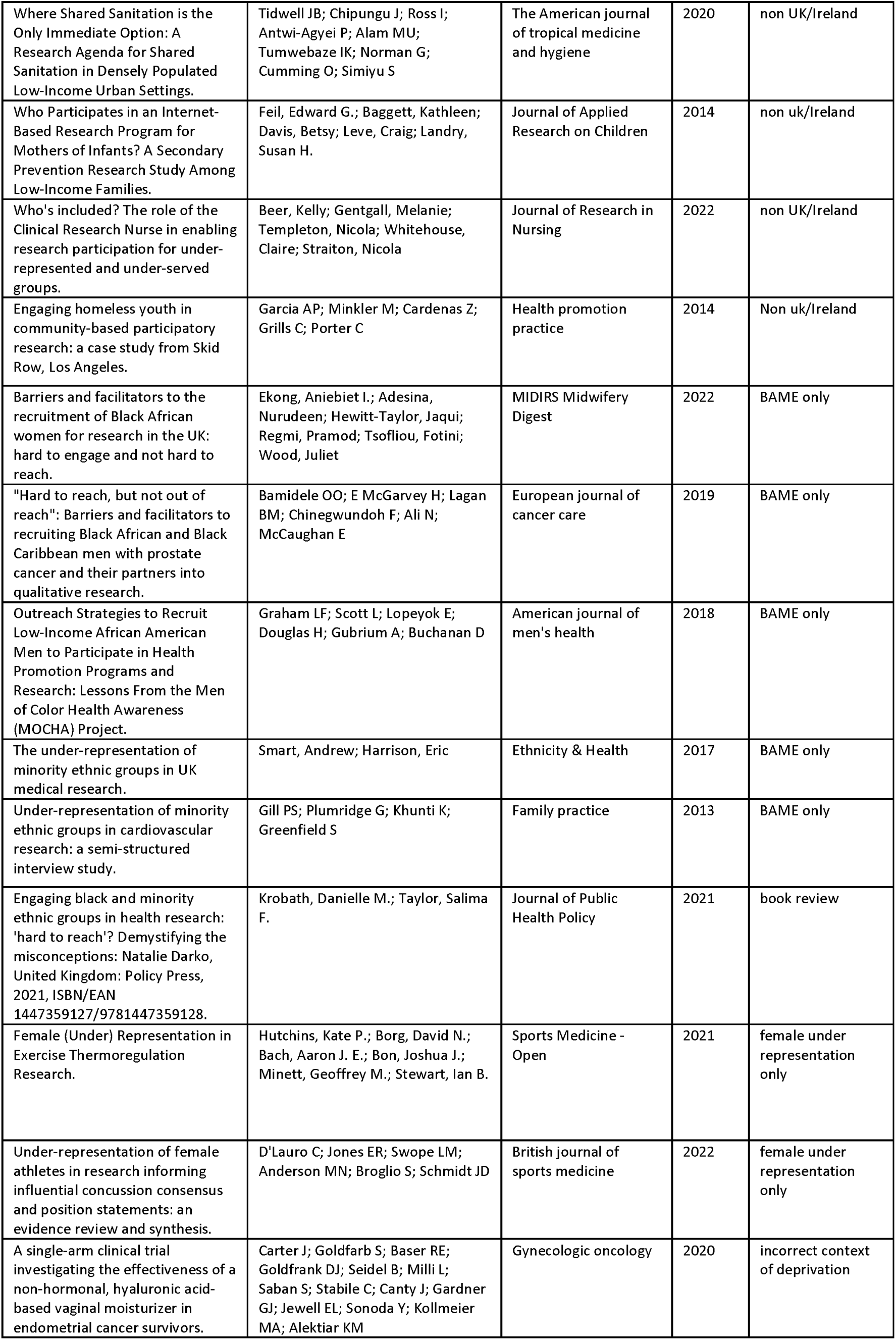
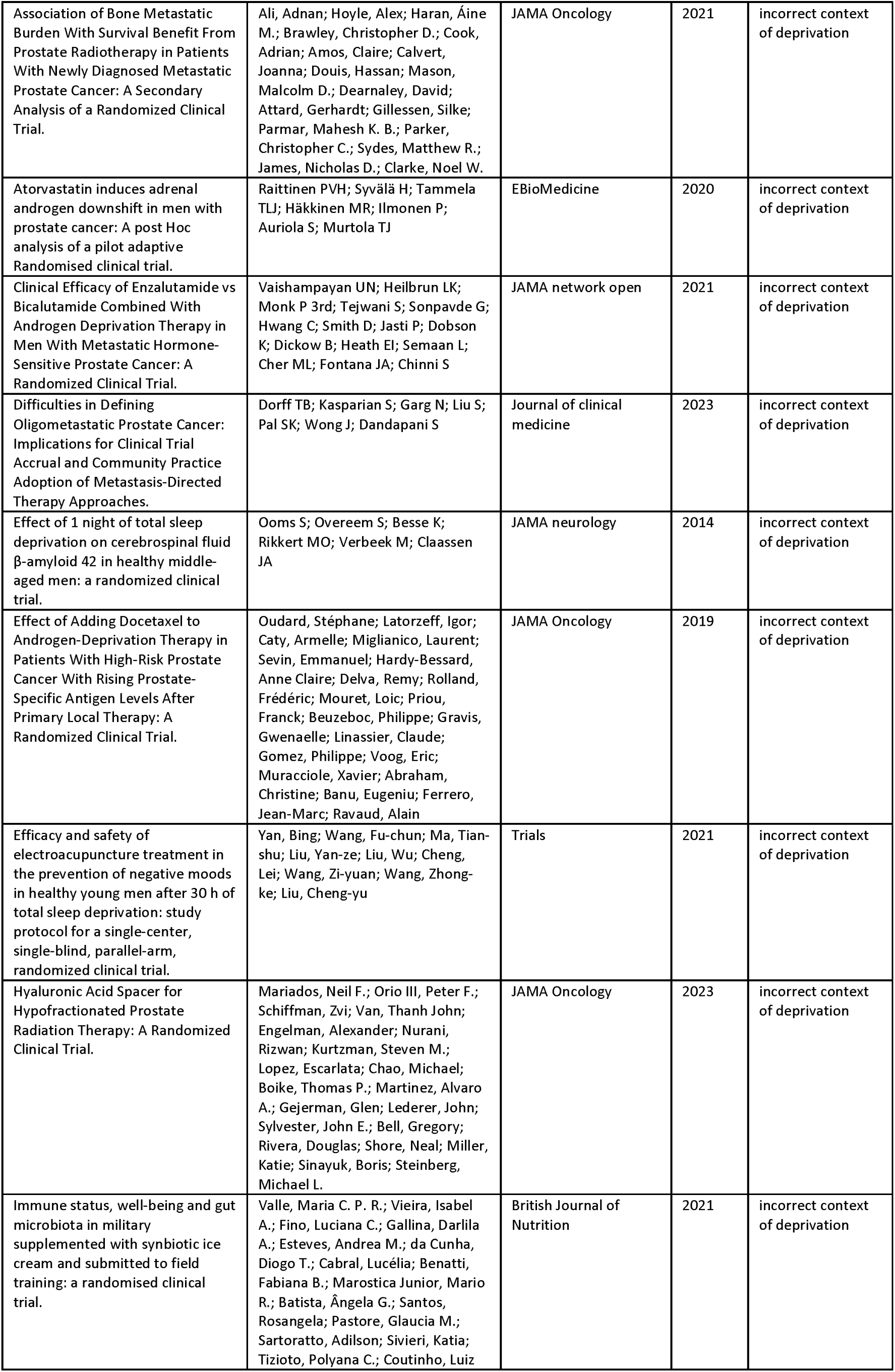
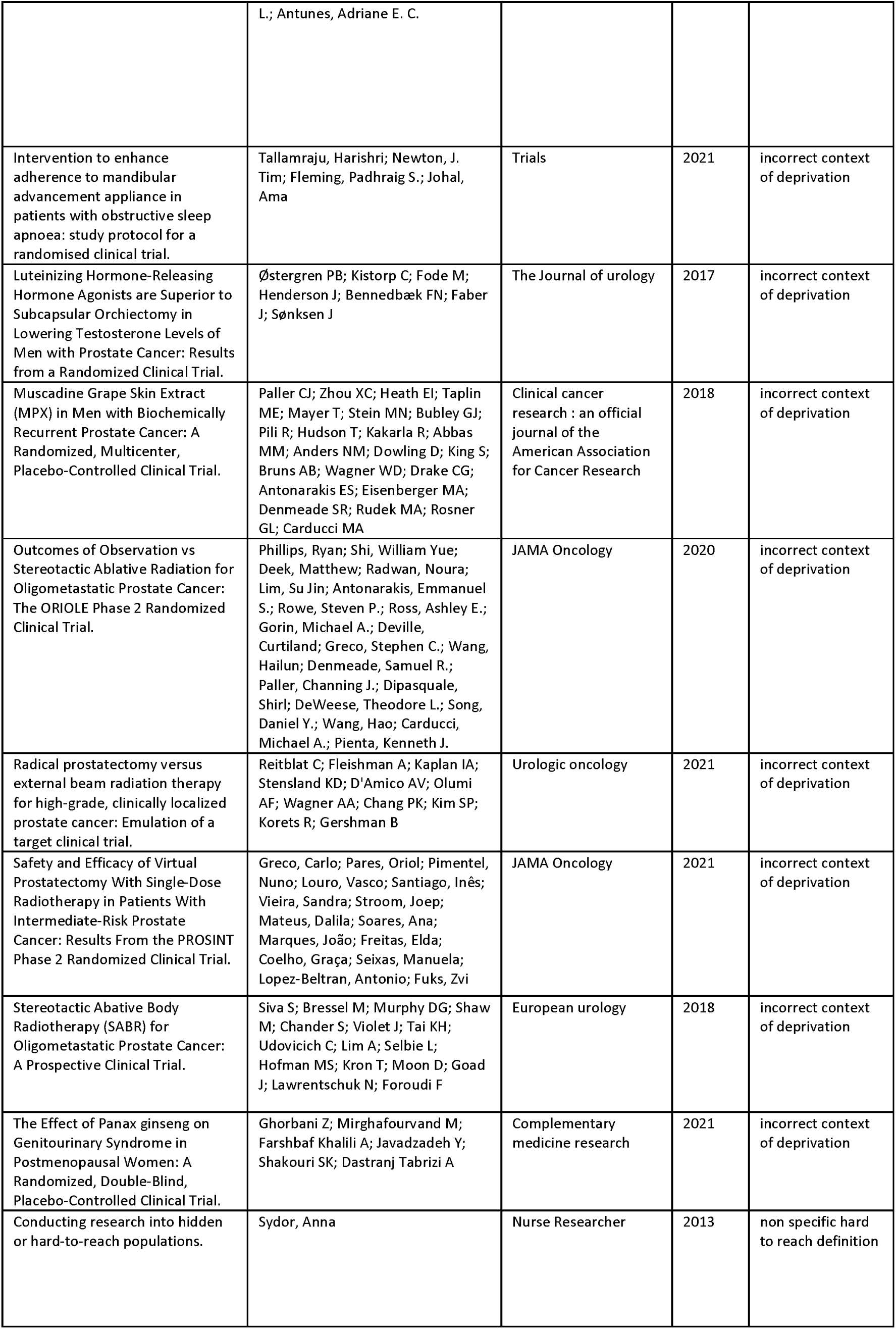
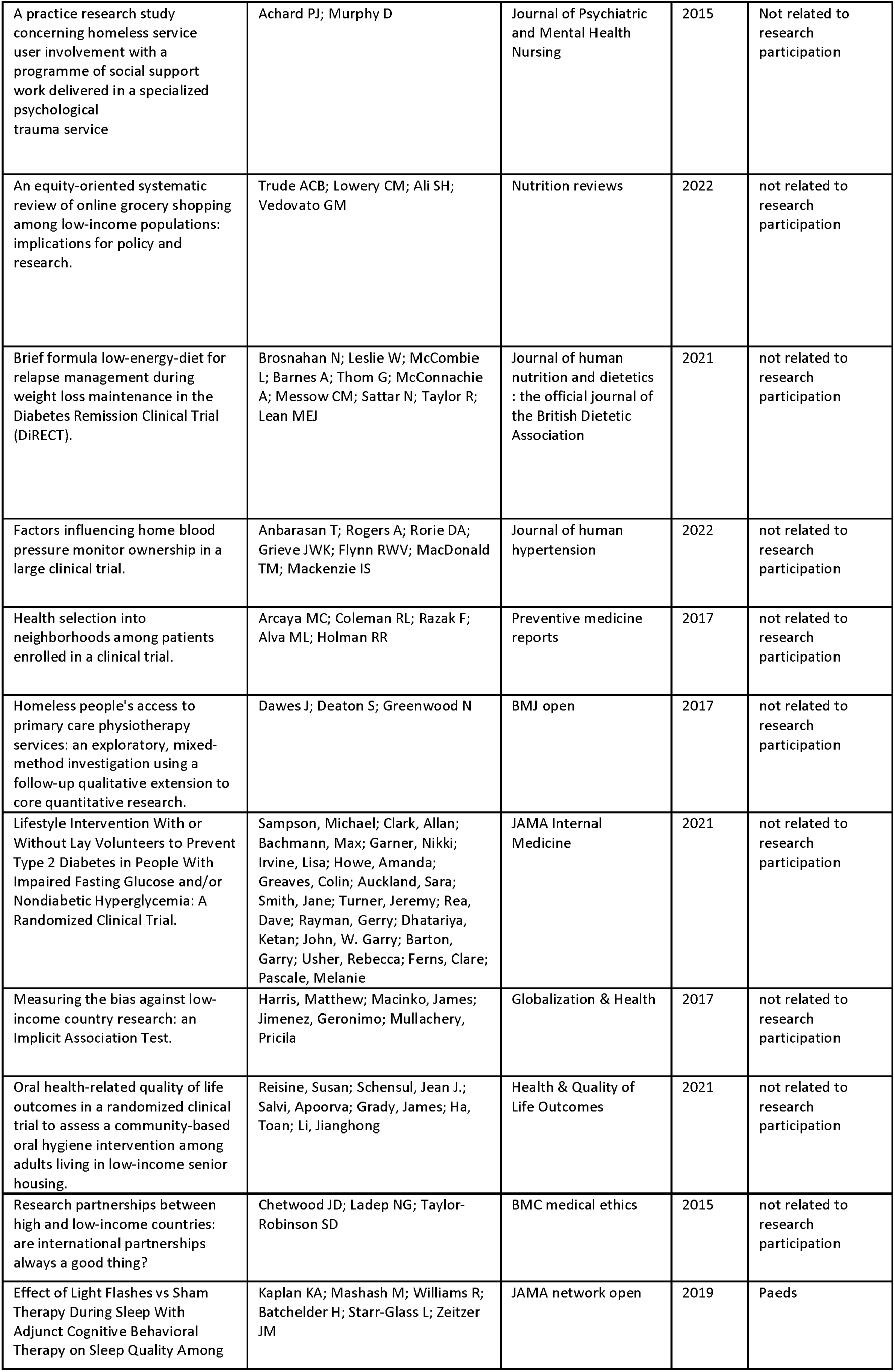
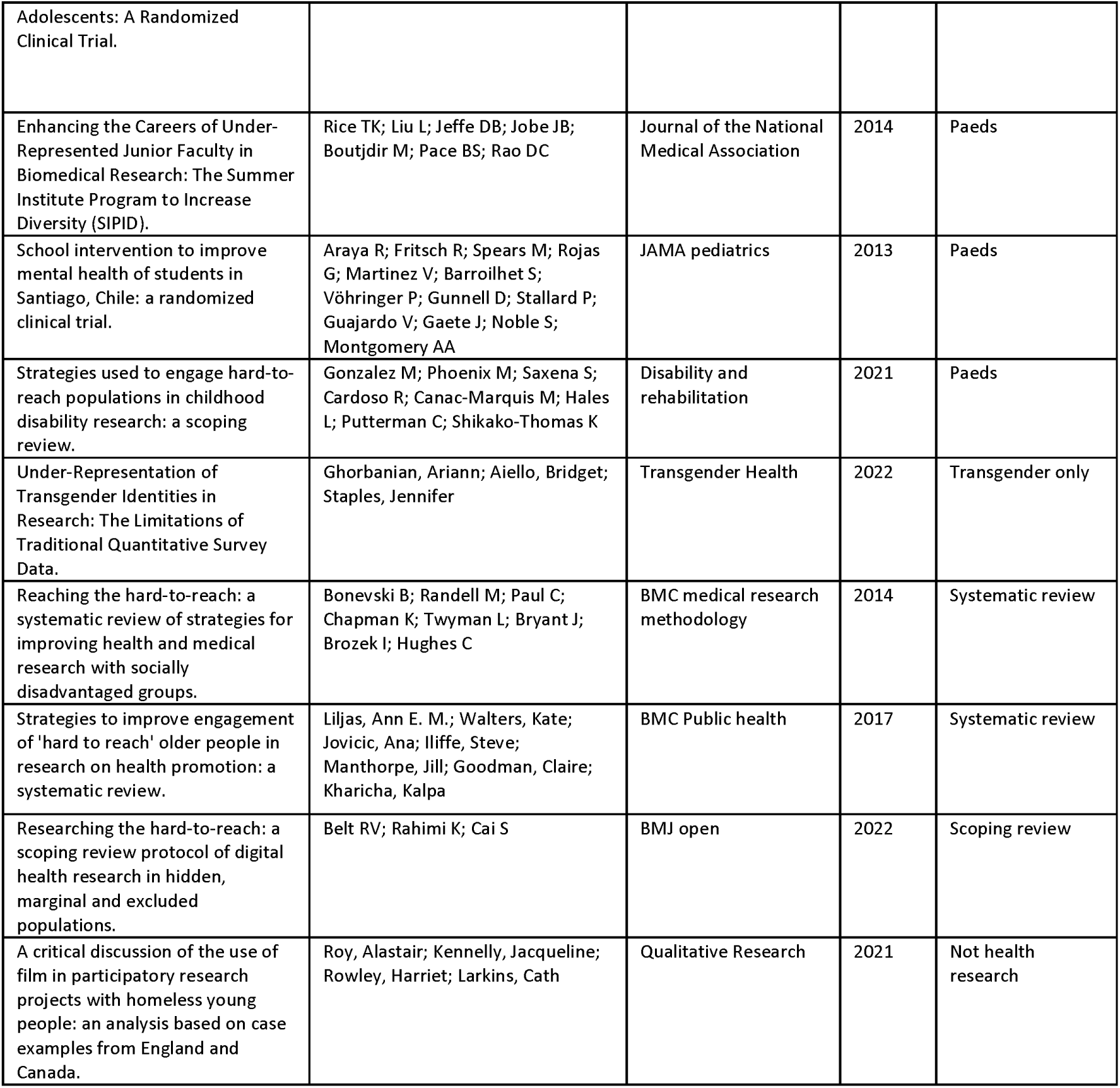
List of excluded papers.

**Appendix 3.**
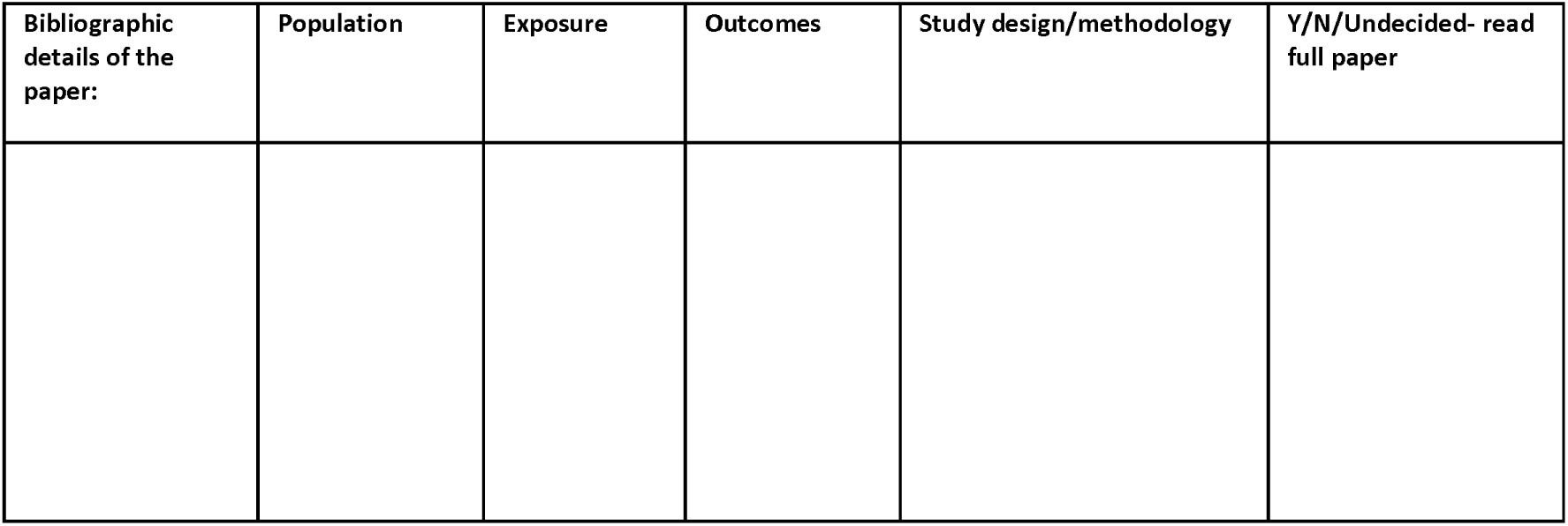
PEOS template.

**Appendix 4.**
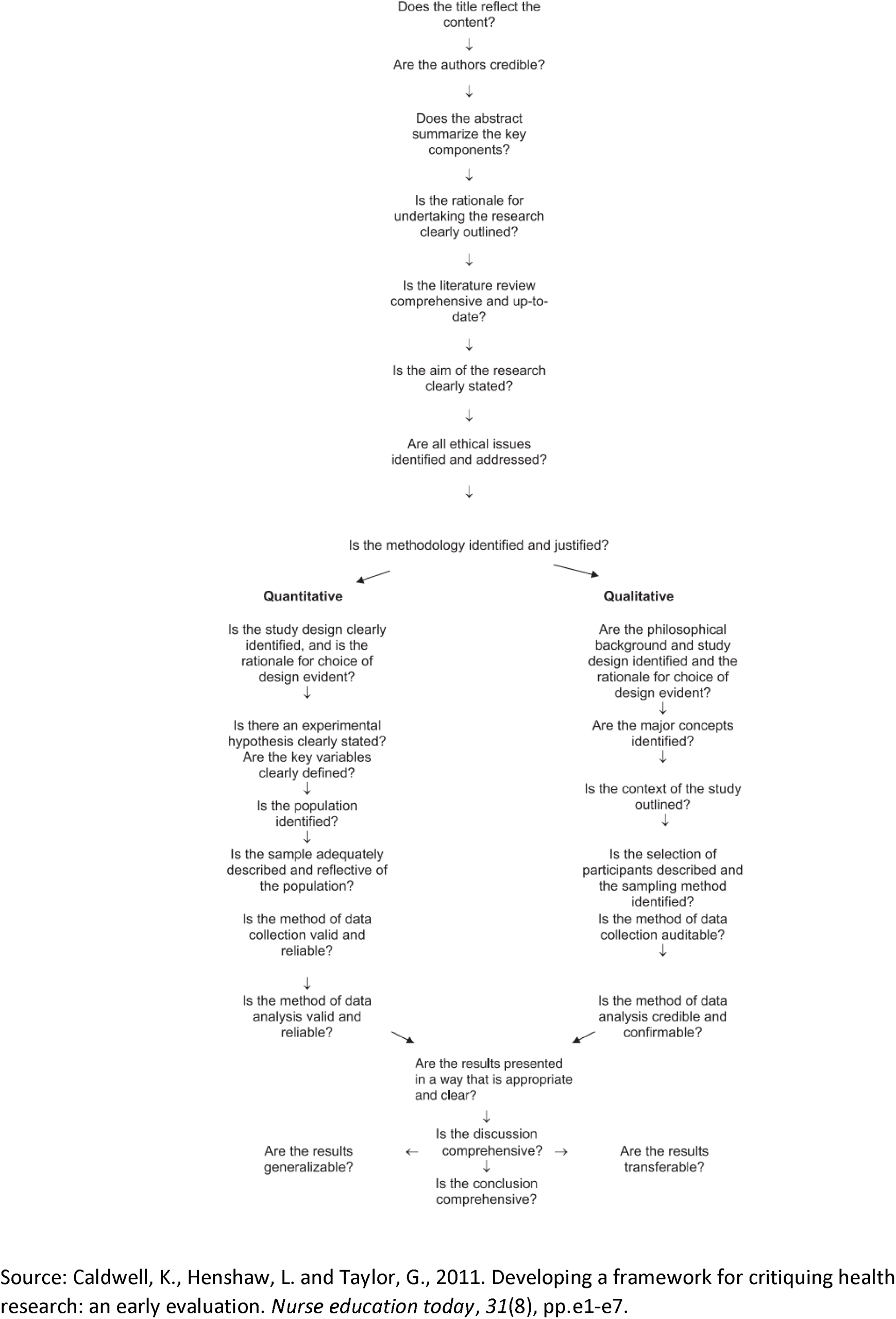
Caldwell framework flow chart

**Appendix 5.** Combined Caldwell Evaluation Table for Overall Numerical Score.

| Q. | Questions for qualitative studies based on the Caldwell framework | Biswell et al, 2021 | Condon et al, 2019 | Condon et al, 2022 | Islam et al, 2021 | O'Reilly-de Brun et al, 2016 | Smyth et al, 2020 | Tully et al, 2021 | Walsh et al, 2022 |
| --- | --- | --- | --- | --- | --- | --- | --- | --- | --- |
|  | Population group | Homeless | GTR | GTR | SE dep | SE dep | GTR | SE dep | GTR & homeless |
| 1 | Does the title reflect the content | 1 | 1 | 2 | 2 | 2 | 1 | 2 | 2 |
| 2 | Are the authors credible (does it state qualifications, who they are and where they work?) | 2 | 1 | 2 | 1 | 1 | 1 | 1 | 1 |
| 3 | Does the abstract summarise the contents | 2 | 1 | 2 | 1 | 1 | 1 | 2 | 1 |
| 4 | Rationale for undertaking the research clearly outlined | 1 | 2 | 2 | 2 | 1 | 2 | 2 | 2 |
| 5 | Is the literature review comprehensive and up-to-date | 2 | 2 | 2 | 2 | 2 | 1 | 2 | 2 |
| 6 | Are the aims clearly stated | 2 | 1 | 2 | 2 | 1 | 1 | 2 | 1 |
| 7 | Ethical issues identified and addressed? | 2 | 2 | 2 | 1 | 2 | 1 | 2 | 2 |
| 8 | Is the methodology identified and justified | 1 | 2 | 2 | 1 | 2 | 1 | 2 | 2 |
| 9 | Are the philosophical background and study design identified and the rationale for choice evident? | 2 | 2 | 2 | 2 | 2 | 1 | 2 | 2 |
| 10 | Are the major concepts identified? | 2 | 2 | 2 | 2 | 2 | 1 | 2 | 2 |
| 11 | Is the context of the study outlined? | 2 | 2 | 2 | 2 | 2 | 1 | 2 | 2 |
| 12 | Is the selection of participants described and sampling method identified? | 2 | 1 | 2 | 1 | 2 | 1 | 2 | 1 |
| 13 | Is the method of data collection auditable? | 1 | 1 | 1 | 1 | 2 | 1 | 2 | 1 |
| 14 | Is the method of data analysis credible and confirmable? | 2 | 1 | 1 | 1 | 2 | 0 | 2 | 1 |
| 15 | Are the results presented in a way that is appropriate and clear | 2 | 1 | 1 | 2 | 2 | 1 | 2 | 1 |
| 16 | Is the discussion comprehensive | 2 | 2 | 2 | 2 | 2 | 1 | 2 | 2 |
| 17 | Is the conclusion comprehensive | 2 | 2 | 1 | 2 | 1 | 2 | 1 | 2 |
| 18 | Are the results transferable | 1 | 1 | 1 | 2 | 2 | 1 | 2 | 1 |
|  | Numerical assessment awarded by author (max score is 36) | 31/36 | 27/36 | 31/36 | 29/36 | 31/36 | 19/36 | 34/36 | 28/36 |

| Q. | Questions for quantitative studies based on the Caldwell framework | Astbury et al, 2020 | Dewhurst et al, 2022 | Quaife et al, 2019 | Rogers et al, 2019 | Vickenson et al, 2021 |
| --- | --- | --- | --- | --- | --- | --- |
|  | Population | SE dep | SE dep | SE dep | SE dep | SE dep |
| 1 | Does the title reflect the content | 2 | 2 | 1 | 1 | 2 |
| 2 | Are the authors credible (does it state qualifications, who they are and where they work?) | 1 | 1 | 1 | 1 | 2 |
| 3 | Does the abstract summarise the contents | 2 | 2 | 2 | 2 | 1 |
| 4 | Rationale for undertaking the research clearly outlined | 2 | 2 | 2 | 2 | 2 |
| 5 | Is the literature review comprehensive and up-to-date | 2 | 2 | 2 | 2 | 2 |
| 6 | Are the aims clearly stated | 1 | 2 | 2 | 2 | 2 |
| 7 | Ethical issues identified and addressed? | 2 | 2 | 2 | 2 | 2 |
| 8 | Is the methodology identified and justified | 2 | 2 | 2 | 2 | 2 |
| 9 | Is the study design clearly identified, and is the rationale for choice of design evident | 2 | 2 | 2 | 2 | 2 |
| 10 | Is there an experimental hypothesis clearly stated and are the key variables clearly defined | 2 | 2 | 2 | 2 | 2 |
| 11 | Is the population identified | 2 | 2 | 2 | 2 | 2 |
| 12 | Is the sample adequately described and reflective of the population | 2 | 2 | 2 | 1 | 2 |
| 13 | Is the method of data collection valid and reliable | 2 | 2 | 2 | 2 | 1 |
| 14 | Is the method of data analysis valid and reliable | 2 | 2 | 2 | 2 | 2 |
| 15 | Are the results presented in a way that is appropriate and clear | 2 | 2 | 2 | 2 | 2 |
| 16 | Is the discussion comprehensive | 2 | 2 | 2 | 2 | 2 |
| 17 | Is the conclusion comprehensive | 2 | 2 | 2 | 2 | 2 |
| 18 | Are the results generalizable | 2 | 2 | 1 | 1 | 2 |
|  | Numerical assessment awarded by author (max score is 36) | 34/36 | 35/36 | 33/36 | 32/36 | 34/36 |

**Appendix 6.**
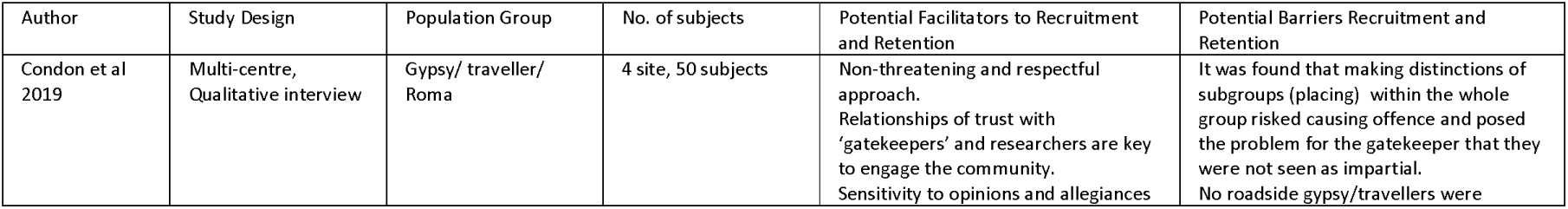
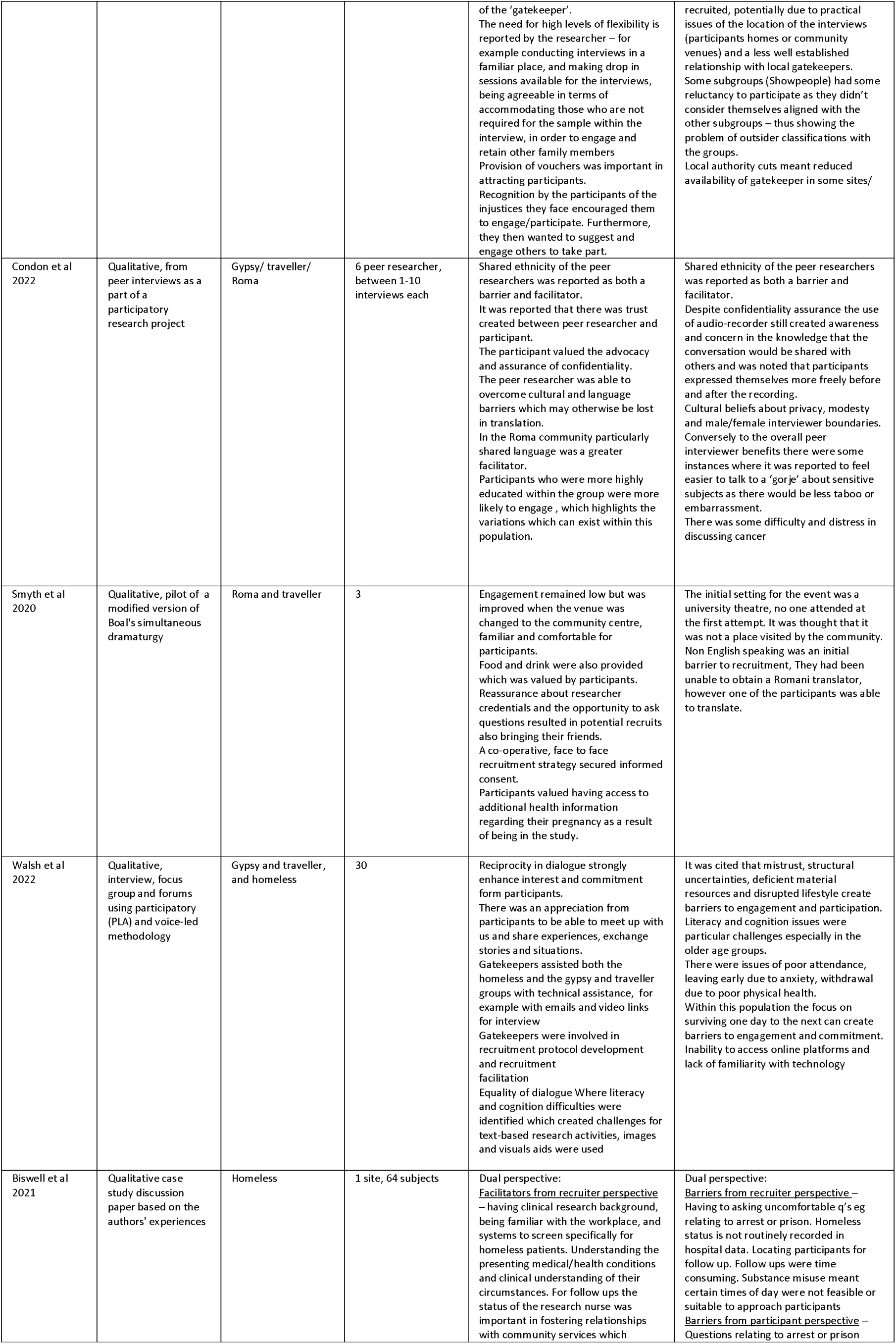
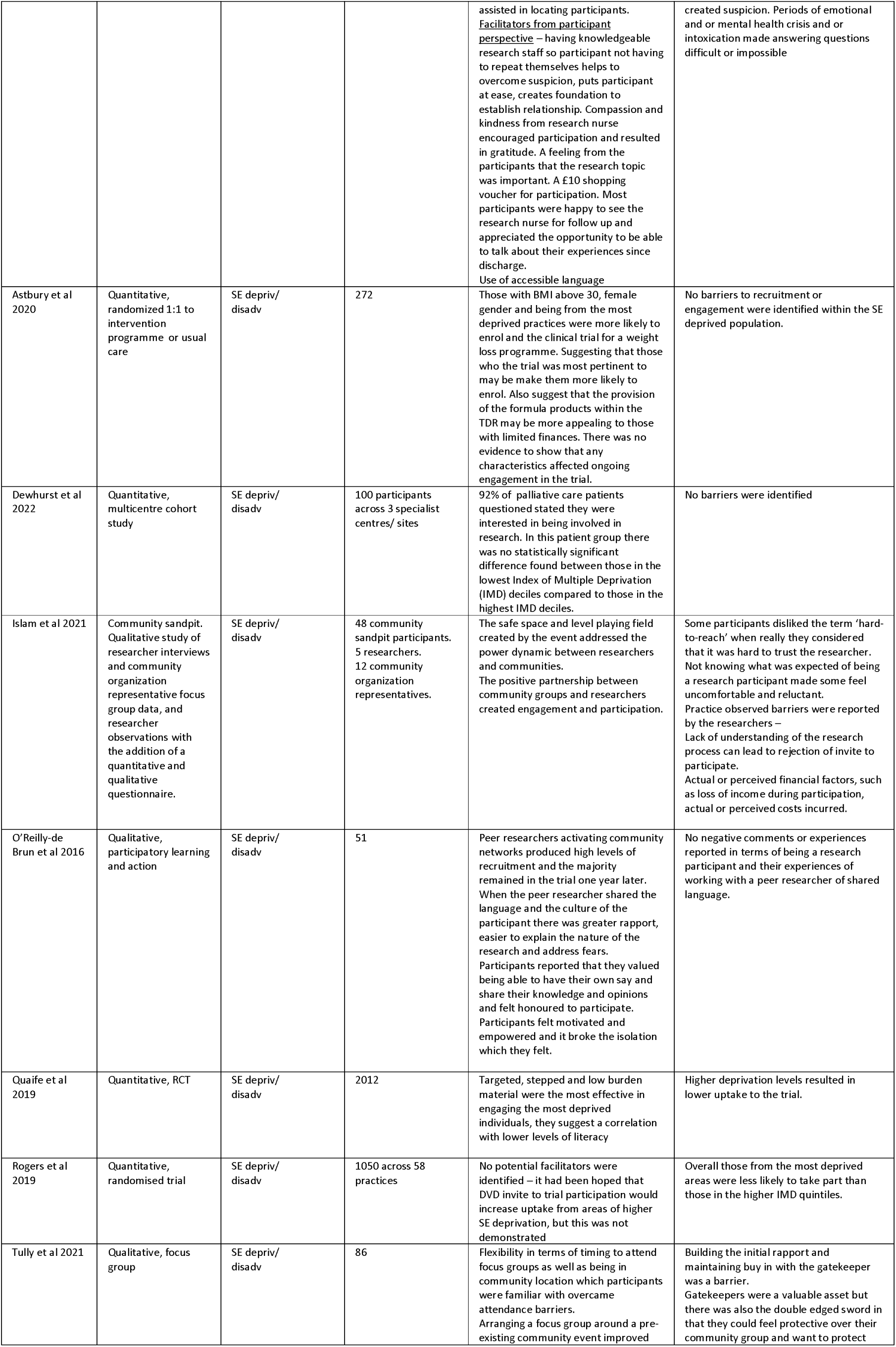
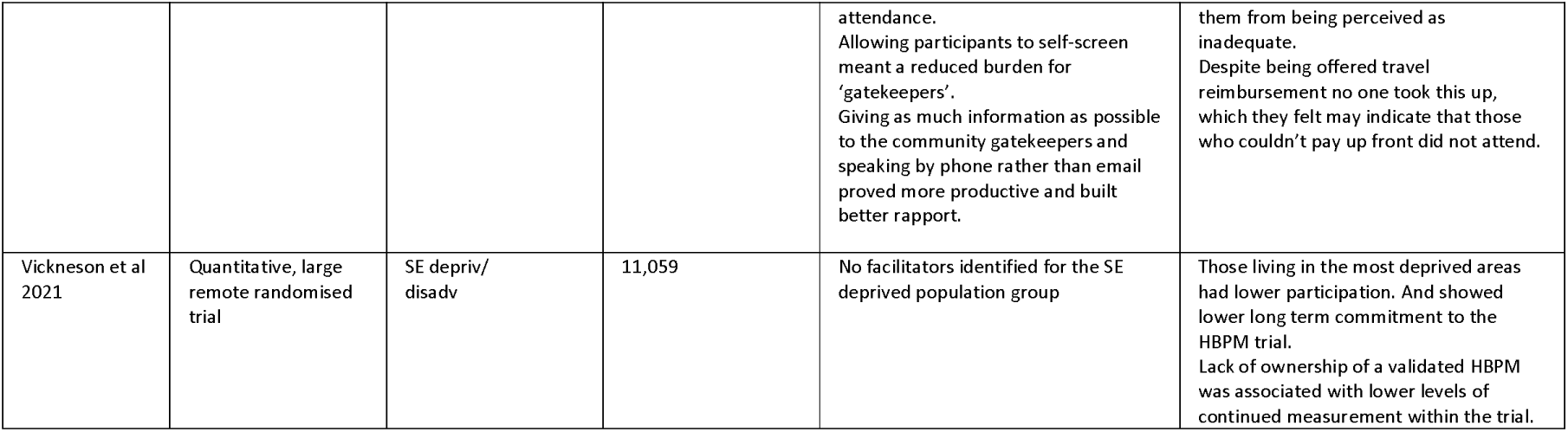
Data extraction of study characteristics.

| Author | Study Design | Population Group | No. of subjects | Potential Facilitators to Recruitment and Retention | Potential Barriers Recruitment and Retention |
| --- | --- | --- | --- | --- | --- |
| Condon et al 2019 | Multi-centre, Qualitative interview | Gypsy/ traveller/ Roma | 4 site, 50 subjects | Non-threatening and respectful approach.<br>Relationships of trust with 'gatekeepers' and researchers are key to engage the community.<br>Sensitivity to opinions and allegiances | It was found that making distinctions of subgroups (placing) within the whole group risked causing offence and posed the problem for the gatekeeper that they were not seen as impartial.<br>No roadside gypsy/travellers were |
|  |  |  |  | of the 'gatekeeper'. The need for high levels of flexibility is reported by the researcher – for example conducting interviews in a familiar place, and making drop in sessions available for the interviews, being agreeable in terms of accommodating those who are not required for the sample within the interview, in order to engage and retain other family members. Provision of vouchers was important in attracting participants. Recognition by the participants of the injustices they face encouraged them to engage/participate. Furthermore, they then wanted to suggest and engage others to take part. | recruited, potentially due to practical issues of the location of the interviews (participants homes or community venues) and a less well established relationship with local gatekeepers. Some subgroups (Showpeople) had some reluctance to participate as they didn't consider themselves aligned with the other subgroups – thus showing the problem of outsider classifications with the groups. Local authority cuts meant reduced availability of gatekeeper in some sites/ |
| Condon et al 2022 | Qualitative, from peer interviews as a part of a participatory research project | Gypsy/ traveller/ Roma | 6 peer researcher, between 1-10 interviews each | Shared ethnicity of the peer researchers was reported as both a barrier and facilitator. It was reported that there was trust created between peer researcher and participant. The participant valued the advocacy and assurance of confidentiality. The peer researcher was able to overcome cultural and language barriers which may otherwise be lost in translation. In the Roma community particularly shared language was a greater facilitator. Participants who were more highly educated within the group were more likely to engage, which highlights the variations which can exist within this population. | Shared ethnicity of the peer researchers was reported as both a barrier and facilitator. Despite confidentiality assurance the use of audio-recorder still created awareness and concern in the knowledge that the conversation would be shared with others and was noted that participants expressed themselves more freely before and after the recording. Cultural beliefs about privacy, modesty and male/female interviewer boundaries. Conversely to the overall peer interviewer benefits there were some instances where it was reported to feel easier to talk to a 'gorje' about sensitive subjects as there would be less taboo or embarrassment. There was some difficulty and distress in discussing cancer |
| Smyth et al 2020 | Qualitative, pilot of a modified version of Boal's simultaneous dramaturgy | Roma and traveller | 3 | Engagement remained low but was improved when the venue was changed to the community centre, familiar and comfortable for participants. Food and drink were also provided which was valued by participants. Reassurance about researcher credentials and the opportunity to ask questions resulted in potential recruits also bringing their friends. A co-operative, face to face recruitment strategy secured informed consent. Participants valued having access to additional health information regarding their pregnancy as a result of being in the study. | The initial setting for the event was a university theatre, no one attended at the first attempt. It was thought that it was not a place visited by the community. Non English speaking was an initial barrier to recruitment, They had been unable to obtain a Romanian translator, however one of the participants was able to translate. |
| Walsh et al 2022 | Qualitative, interview, focus group and forums using participatory (PLA) and voice-led methodology | Gypsy and traveller, and homeless | 30 | Reciprocity in dialogue strongly enhance interest and commitment from participants. There was an appreciation from participants to be able to meet up with us and share experiences, exchange stories and situations. Gatekeepers assisted both the homeless and the gypsy and traveller groups with technical assistance, for example with emails and video links for interview. Gatekeepers were involved in recruitment protocol development and recruitment facilitation. Equality of dialogue Where literacy and cognition difficulties were identified which created challenges for text-based research activities, images and visual aids were used | It was cited that mistrust, structural uncertainties, deficient material resources and disrupted lifestyle create barriers to engagement and participation. Literacy and cognition issues were particular challenges especially in the older age groups. There were issues of poor attendance, leaving early due to anxiety, withdrawal due to poor physical health. With in this population the focus on surviving one day to the next can create barriers to engagement and commitment. Inability to access online platforms and lack of familiarity with technology |
| Biswell et al 2021 | Qualitative case study discussion paper based on the authors' experiences | Homeless | 1 site, 64 subjects | Dual perspective: <u>Facilitators from recruiter perspective</u> – having clinical research background, being familiar with the workplace, and systems to screen specifically for homeless patients. Understanding the presenting medical/health conditions and clinical understanding of their circumstances. For follow ups the status of the research nurse was important in fostering relationships with community services which | Dual perspective: <u>Barriers from recruiter perspective</u> – Having to asking uncomfortable questions relating to arrest or prison. Homeless status is not routinely recorded in hospital data. Locating participants for follow up. Follow ups were time consuming. Substance misuse meant certain times of day were not feasible or suitable to approach participants. <u>Barriers from participant perspective</u> – Questions relating to arrest or prison |
|  |  |  |  | assisted in locating participants. <u>Facilitators from participant perspective</u> – having knowledgeable research staff so participant not having to repeat themselves helps to overcome suspicion, puts participant at ease, creates foundation to establish relationship. Compassion and kindness from research nurse encouraged participation and resulted in gratitude. A feeling from the participants that the research topic was important. A £10 shopping voucher for participation. Most participants were happy to see the research nurse for follow up and appreciated the opportunity to be able to talk about their experiences since discharge.<br>Use of accessible language | created suspicion. Periods of emotional and or mental health crisis and or intoxication made answering questions difficult or impossible |
| Astbury et al 2020 | Quantitative, randomized 1:1 to intervention programme or usual care | SE depriv/ disadv | 272 | Those with BMI above 30, female gender and being from the most deprived practices were more likely to enrol and the clinical trial for a weight loss programme. Suggesting that those who the trial was most pertinent to may be make them more likely to enrol. Also suggest that the provision of the formula products within the TDR may be more appealing to those with limited finances. There was no evidence to show that any characteristics affected ongoing engagement in the trial. | No barriers to recruitment or engagement were identified within the SE deprived population. |
| Dewhurst et al 2022 | Quantitative, multicentre cohort study | SE depriv/ disadv | 100 participants across 3 specialist centres/sites | 92% of palliative care patients questioned stated they were interested in being involved in research. In this patient group there was no statistically significant difference found between those in the lowest Index of Multiple Deprivation (IMD) deciles compared to those in the highest IMD deciles. | No barriers were identified |
| Islam et al 2021 | Community sandpit. Qualitative study of researcher interviews and community organization representative focus group data, and researcher observations with the addition of a quantitative and qualitative questionnaire. | SE depriv/ disadv | 48 community sandpit participants. 5 researchers. 12 community organization representatives. | The safe space and level playing field created by the event addressed the power dynamic between researchers and communities. The positive partnership between community groups and researchers created engagement and participation. | Some participants disliked the term 'hard-to-reach' when really they considered that it was hard to trust the researcher. Not knowing what was expected of being a research participant made some feel uncomfortable and reluctant. Practice observed barriers were reported by the researchers – Lack of understanding of the research process can lead to rejection of invite to participate. Actual or perceived financial factors, such as loss of income during participation, actual or perceived costs incurred. |
| O'Reilly-de Brun et al 2016 | Qualitative, participatory learning and action | SE depriv/ disadv | 51 | Peer researchers activating community networks produced high levels of recruitment and the majority remained in the trial one year later. When the peer researcher shared the language and the culture of the participant there was greater rapport, easier to explain the nature of the research and address fears. Participants reported that they valued being able to have their own say and share their knowledge and opinions and felt honoured to participate. Participants felt motivated and empowered and it broke the isolation which they felt. | No negative comments or experiences reported in terms of being a research participant and their experiences of working with a peer researcher of shared language. |
| Quaife et al 2019 | Quantitative, RCT | SE depriv/ disadv | 2012 | Targeted, stepped and low burden material were the most effective in engaging the most deprived individuals, they suggest a correlation with lower levels of literacy | Higher deprivation levels resulted in lower uptake to the trial. |
| Rogers et al 2019 | Quantitative, randomised trial | SE depriv/ disadv | 1050 across 58 practices | No potential facilitators were identified – it had been hoped that DVD invite to trial participation would increase uptake from areas of higher SE deprivation, but this was not demonstrated | Overall those from the most deprived areas were less likely to take part than those in the higher IMD quintiles. |
| Tully et al 2021 | Qualitative, focus group | SE depriv/ disadv | 86 | Flexibility in terms of timing to attend focus groups as well as being in community location which participants were familiar with overcame attendance barriers. Arranging a focus group around a pre-existing community event improved | Building the initial rapport and maintaining buy in with the gatekeeper was a barrier. Gatekeepers were a valuable asset but there was also the double edged sword in that they could feel protective over their community group and want to protect |
|  |  |  |  | attendance.<br>Allowing participants to self-screen meant a reduced burden for 'gatekeepers'.<br>Giving as much information as possible to the community gatekeepers and speaking by phone rather than email proved more productive and built better rapport. | them from being perceived as inadequate.<br>Despite being offered travel reimbursement no one took this up, which they felt may indicate that those who couldn't pay up front did not attend. |
| Vickneson et al 2021 | Quantitative, large remote randomised trial | SE depriv/ disadv | 11,059 | No facilitators identified for the SE deprived population group | Those living in the most deprived areas had lower participation. And showed lower long term commitment to the HBPM trial.<br>Lack of ownership of a validated HBPM was associated with lower levels of continued measurement within the trial. |

**Appendix 7.** A tabulation of the themes of barriers and facilitators for each population group.

|  | Facilitators | Barriers |
| --- | --- | --- |
| Gypsy and traveller | Non-threatening and respectful approach.<br>Relationships of trust with community representatives/leaders, researchers and participants | Generalising of subgroups within the overarching group, leading to potential of stigma, causing offence and lack of identification.<br>Practical issues of the location of |
|  | • Sensitivity to opinions and allegiances of the 'gatekeeper'.<br>• Gatekeepers provided technical assistance, eg emails and video links.<br>• Flexibility: conducting interviews in a familiar place, drop in sessions available for the interviews, being accommodating with sample to engage other family members<br>• Provision of vouchers<br>• Provision of food and drink<br>• Recognition of the participants injustices.<br>• Opportunity to suggest and engage others to take part.<br>• Shared ethnicity of peer researchers<br>• Advocacy and assurance of confidentiality.<br>• Researcher ability to overcome cultural and language barriers.<br>• Reciprocal dialogue<br>• Allowances for variations in education and literacy<br>• Familiarity and comfort of the venue<br>• Reassurance about researcher credentials.<br>• Opportunity to ask questions<br>• Co-operative, face to face recruitment strategy.<br>• Participant access to additional information regarding their health.<br>• Opportunity to meet up with others and share experiences, exchange stories and situations. | the interviews with no permanent address<br>• Less well established relationship with local gatekeepers.<br>• Reduced availability of gatekeeper due to local authority cutbacks in some sites<br>• Shared ethnicity of the peer researchers<br>• The use of audio-recorder raised awareness that the conversation would be shared and caused hesitancy<br>• Cultural beliefs about privacy, modesty and male/female interviewer boundaries.<br>• Some instances it was reported as easier to talk to a an 'outsider' about sensitive subjects as less taboo or embarrassment.<br>• Difficulty and distress in discussing cancer<br>• Venue unknown or unfamiliar to the community.<br>• Non-English speaking recruiters<br>• Mistrust, structural uncertainties, deficient material resources and disrupted lifestyle<br>• Literacy issues<br>• Participant anxiety and/or poor physical health<br>• Inability to access online platforms could mean that some were unable to take part |
| SE deprived | • Those who the trial was most pertinent more likely to enrol for example in obese, deprived population and also in palliative care were strong drivers to participate<br>• Provision of medical/health trial related products seemed more appealing to those with limited finances.<br>• A safe space, level playing field, balanced power dynamic between researchers and communities.<br>• Positive partnership between community groups and researchers<br>• Peer researchers activating community networks<br>• Shared the language and the culture of the peer researcher and participant created rapport and engagement<br>• Participants being able to have their own say, share their knowledge and | • Stigma around term and label of 'hard-to-reach'<br>• Hard to trust the researcher.<br>• Not knowing what was expected of being a research participant.<br>• Lack of understanding of the research process can lead to rejection of invite to participate.<br>• Actual or perceived financial factors, such as loss of income during participation, actual or perceived costs incurred<br>• Higher deprivation levels resulted in lower uptake to the trial and reduced commitment to trial<br>• Challenges around building initial rapport and maintaining relationship with community gatekeeper<br>• Gatekeepers could feel protective over their community group and |
|  | opinions.<br>• Sense of honour to participate.<br>• Participant feeling motivated and empowered<br>• Reduction in feelings of isolation<br>• Targeted, stepped approach and low burden trial materials<br>• Flexibility in timings for focus groups, arranging it around a pre-existing community event<br>• Familiar community location.<br>• Allowing participants to self-screen for approach<br>• Enabling well informed community gatekeepers and liaising with them by phone rather than email | want to protect them from being labelled or perceived as inadequate<br>• Not possessing the necessary requirement for participating in the trial |
| Homeless | <u>Recruiter perspective</u><br>• Clinical research background, familiarity with the workplace, and systems, understanding medical/health conditions and circumstances.<br>• Status of the research nurse was important in fostering relationships with community services which assisted in locating participants.<br><u>Participant perspective</u><br>• Knowledgeable research staff meant not having to repeat themselves and helps to overcome suspicion, puts participant at ease, creates foundation to establish relationship.<br>• A feeling from the participants that the research topic was important.<br>• Able to address the unmet needs of homeless patients on the ward by knowing where and how to access resources within hospital which meant being able to provide clothing, toiletries and wash dirty clothes,<br>• This compassionate and kindness on the part of the research nurse, encouraged participation and patients expressed gratitude for this help<br>• Provision of a voucher for participation.<br>• The opportunity to be able to talk about their experiences, exchange stories and situations<br>• Gatekeepers helped to provide technical assistance<br>• Equality of dialogue and use of accessible language<br>• Use of visual materials rather than text | <u>Recruiter perspective</u><br>• Having to asking uncomfortable q's eg relating to arrest or prison.<br>• Homeless status not routinely recorded in hospital data.<br>• Locating participants for follow up.<br>• Follow ups were time consuming.<br>• Substance misuse meant certain times of day were not feasible or suitable to approach participants<br><u>Participant perspective</u><br>• Questions relating to arrest or prison created suspicion.<br>• Periods of emotional and or mental health crisis and or intoxication made answering questions difficult or impossible<br>• Mistrust, structural uncertainties, deficient material resources and disrupted lifestyle<br>• Literacy issues<br>• Anxiety and/or poor physical health.<br>• The focus on surviving one day to the next is often priority<br>• Inability to access online platforms could mean that some were unable to take part |

## Notes

### Competing Interest Statement

The authors have declared no competing interest.

### Funding Statement

This study did not receive any funding

## References

Aldridge, R.W., Story, A., Hwang, S.W., Nordentoft, M., Luchenski, S.A., Hartwell, G., Tweed, E.J., Lewer, D., Katikireddi, S.V. and Hayward, A.C., 2018. Morbidity and mortality in homeless individuals, prisoners, sex workers, and individuals with substance use disorders in high-income countries: a systematic review and meta-analysis. The Lancet, [online] 391(10117), pp.241–250. available at: <10.1016/S0140-6736(17)31869-X> [accessed 10/05/2023]

Al-Shakarchi, N.J., Evans, H., Luchenski, S.A., Story, A. and Banerjee, A., 2020. Cardiovascular disease in homeless versus housed individuals: a systematic review of observational and interventional studies. Heart, [online] 106(19), pp.1483–1488. available at: <10.1136/heartjnl-2020-316706> [accessed 10/05/2023]

Aromataris, E. and Pearson, A., 2014. The systematic review: an overview. AJN The American Journal of Nursing, [online] 114(3), pp.53–58. available at: <10.1097/01.naj.0000444496.24228.2c> [accessed 15/07/2023]

Astbury, N.M., Tudor, K., Aveyard, P. and Jebb, S.A., 2020. Heterogeneity in the uptake, attendance, and outcomes in a clinical trial of a total diet replacement weight loss programme. BMC medicine, [online] 18(1), pp.1–9. available at: <10.1186/s12916-020-01547-4> [accessed 17/05/2023]

Belt, R.V., Rahimi, K. and Cai, S., 2022. Researching the hard-to-reach: a scoping review protocol of digital health research in hidden, marginal and excluded populations. BMJ open, [online] 12(9), p.e061361. available at: <10.1136/bmjopen-2022-061361>[accessed on 17/07/2023]

Bettany-Saltikov, J. and McSherry R (2016). How to do a Systematic Literature Review in Nursing: A step-by-step guide. 2nd Ed. Open University Press: London

Biswell R E., Clark, M., Tinelli, M., Manthorpe, G., Neale, J., Whiteford, M. and Cornes, M., 2021. Beyond clinical trials: Extending the role of the clinical research nurse into social care and homeless research. Journal of Clinical Nursing. [online] available at: <10.1111/jocn.15911> [accessed 17/05/2023]

Bonevski, B., Randell, M., Paul, C., Chapman, K., Twyman, L., Bryant, J., Brozek, I. and Hughes, C., 2014. Reaching the hard-to-reach: a systematic review of strategies for improving health and medical research with socially disadvantaged groups. BMC medical research methodology, [online] 14, pp.1–29. available at: <10.1186/1471-2288-14-42> [accessed 11/08/2023]

Bowen, M., Marwick, S., Marshall, T., Saunders, K., Burwood, S., Yahyouche, A., Stewart, D. and Paudyal, V., 2019. Multimorbidity and emergency department visits by a homeless population: a database study in specialist general practice. British Journal of General Practice, [online] 69(685), pp.e515–e525. available at: <https://bjgp.org/content/bjgp/69/685/e515.full.pdf> [accessed 11/08/2023]

Bower, P., Grigoroglou, C., Anselmi, L., Kontopantelis, E., Sutton, M., Ashworth, M., Evans, P., Lock, S., Smye, S. and Abel, K., 2020. Is health research undertaken where the burden of disease is greatest? Observational study of geographical inequalities in recruitment to research in England 2013–2018. BMC medicine, [online] 18, pp.1–11. available at: <https://bmcmedicine.biomedcentral.com/articles/10.1186/s12916-020-01555-4> [accessed 24/07/2023]

Brown, P. and Scullion, L., 2010. ‘Doing research ‘with Gypsy–Travellers in England: reflections on experience and practice. Community Development Journal, [online] 45(2), pp.169–185. available at: <10.1093/cdj/bsp008> [accessed 24/07/2023]

Caldwell, K., Henshaw, L. and Taylor, G., 2011. Developing a framework for critiquing health research: an early evaluation. Nurse education today, [online] 31(8), pp.e1–e7. available at: <10.1016/j.nedt.2010.11.025> [accessed 20/06/2023]

Cancer Research UK (2020) Cancer in the UK 2020: Socio-economic deprivation. [online] available at: <https://www.cancerresearchuk.org/sites/default/files/cancer_inequalities_in_the_uk.pdf> [accessed on 23/06/2023]

Condon, L., Bedford, H., Ireland, L., Kerr, S., Mytton, J., Richardson, Z. and Jackson, C., 2019. Engaging gypsy, Roma, and traveller communities in research: Maximizing opportunities and overcoming challenges. Qualitative Health Research, [online] 29(9), pp.1324–1333. [online] available at: <10.1177/1049732318813558> [accessed 11/08/2023]

Condon, L., Curejova, J., Morgan, D.L., Miles, G. and Fenlon, D., 2021 (a). Knowledge and experience of cancer prevention and screening among Gypsies, Roma and Travellers: a participatory qualitative study. BMC Public Health, [online] 21(1), pp.1–11. available at: <10.1186/s12889-021-10390-y> [accessed 19/05/2023]

Condon, L., Curejova, J., Leeanne Morgan, D. and Fenlon, D., 2021 (b). Cancer diagnosis, treatment and care: A qualitative study of the experiences and health service use of Roma, Gypsies and Travellers. European Journal of Cancer Care,130(5), p.e13439.

Condon L, Curejova J, Leeanne Morgan D, Miles G, Barry D, Fenlon D. (2022). Public involvement in participatory research: the experiences of peer interviewers from Roma, Gypsy and Traveller communities. Nurse Researcher. [online] 2022 Mar 10;30(1):17–23 available at: <10.7748/nr.2022.e1818<otherinfo>> [accessed 17/05/2023]

Cook, B., Wayne, G.F., Valentine, A., Lessios, A. and Yeh, E., 2013. Revisiting the evidence on health and health care disparities among the Roma: a systematic review 2003– 2012. International journal of public health,158, pp.885–911.

Centre for Reviews and Dissemination (2009). Systematic Reviews. CRD’s guidance for undertaking reviews in health care. CRD: University of York. [online] Available at: <https://www.york.ac.uk/media/crd/Systematic_Reviews.pdf> [accessed: 19/05/2023]

Davies, A., 2019. Carrying out systematic literature reviews: an introduction. British Journal of Nursing, [online] 28(15), pp.1008–1014. available at: <10.12968/bjon.2019.28.15.1008> [accessed 15/07/2023]

De los Santos, O.E.M., Alcantar, G.M.L.R. and de Hoyos Bermea, A., 2022. Systematic Reviews And Their Epistemological Foundations: A Narrative Review Of The Literature. Journal of positive school psychology, [online] 6(8), pp.10298–10312. Available at: <http://mail.journalppw.com/index.php/jpsp/article/view/12669/8219> [accessed 15/07/2023]

Department of Health, 2012. Long Term Conditions Compendium of Information. Third Edition. [online] available at: <https://assets.publishing.service.gov.uk/government/uploads/system/uploads/attachment_data/file/216528/dh_134486.pdf> [accessed 19/06/2023]

Dewhurst, F., Wakefield, D., Elverson, J., McConnell, R., Bryan, C., Spriggs, H., Atkinson, K. and Frew, K., 2022. Palliative care inpatients favour research participation irrespective of prognosis, performance or socioeconomic status: multicentre cohort study. BMJ Supportive & Palliative Care. [online] available at: <10.1136/spcare-2022-004037> [accessed 17/05/2023]

Donnelly, D.W. and Gavin, A., 2011. Socio-economic inequalities in cancer incidence–the choice of deprivation measure matters. Cancer epidemiology, [online] 35(6), pp.e55–e61. available at: <10.1016/j.canep.2011.06.002> [accessed 19/05/2023]

Ekezie, W., Czyznikowska, B.M., Rohit, S., Harrison, J., Miah, N., Campbell-Morris, P. and Khunti, K., 2021. The views of ethnic minority and vulnerable communities towards participation in COVID-19 vaccine trials. Journal of Public Health, [online] 43(2), pp.e258–e260. available at: <10.1093/pubmed/fdaa196> [accessed 15/07/2023]

Erves, J.C., Mayo-Gamble, T.L., Malin-Fair, A., Boyer, A., Joosten, Y., Vaughn, Y.C., Sherden, L., Luther, P., Miller, S. and Wilkins, C.H., 2017. Needs, priorities, and recommendations for engaging underrepresented populations in clinical research: a community perspective. Journal of community health, [online] 42, pp.472–480. available at: <10.1007/s10900-016-0279-2> [accessed 10/05/2023]

Gough, D., Thomas, J., & Oliver, S. (2019). Clarifying differences between reviews within evidence ecosystems. Systematic Reviews, [online] 8(1), 170. available at: <10.1186/s13643-019-1089-2> [accessed 11/08/2023]

Gov.uk (2020). People living in deprived neighbourhoods [online] available at: <https://www.ethnicity-facts-figures.service.gov.uk/uk-population-by-ethnicity/demographics/people-living-in-deprived-neighbourhoods/2.1> [accessed 15/07/2023]

Gov.uk (2021). Policy paper: Saving and Improving Lives: The Future of UK Clinical Research Delivery [online] available at:< https://www.gov.uk/government/publications/the-future-of-uk-clinical-research-delivery/saving-and-improving-lives-the-future-of-uk-clinical-research-delivery#our-vision-for-uk-clinical-research-delivery> [accessed 24/07/2023]

Harden A, Sheridan K, McKeown A, Dan-Ogosi I, Bagnall AM (2015) Evidence Review of Barriers to, and Facilitators of, Community Engagement Approaches and Practices in the UK. London: Institute for Health and Human Development, University of East London. [online] available at: <https://www.nice.org.uk/guidance/ng44/evidence/evidence-review-4-map-of-uk-literature-pdf-2368403680> [accessed 30/07/2023]

Harkins, C., Shaw, R., Gillies, M., Sloan, H., MacIntyre, K., Scoular, A., Morrison, C., MacKay, F., Cunningham, H., Docherty, P. and MacIntyre, P., 2010. Overcoming barriers to engaging socio-economically disadvantaged populations in CHD primary prevention: a qualitative study. BMC public health, [online] 10, pp.1–7. Available at < https://link.springer.com/article/10.1186/1471-2458-10-391> [accessed 30/07/2023]

Hunn, A., 2013. Survey of the general public: attitudes towards health research 2013. Government paper. [online] available at: <https://s3.eu-west-2.amazonaws.com/www.hra.nhs.uk/media/documents/survey-general-public-attitudes-towards-health-research.pdf> [accessed 11/08/2023]

Islam, S., Joseph, O., Chaudry, A., Forde, D., Keane, A., Wilson, C., Begum, N., Parsons, S., Grey, T., Holmes, L. and Starling, B., 2021. “We are not hard to reach, but we may find it hard to trust”…. Involving and engaging ‘seldom listened to ‘community voices in clinical translational health research: a social innovation approach. Research Involvement and Engagement, [online] 7(1), p.46. available at: <10.1186/s40900-021-00292-z> [accessed 17/05/2023]

Kelleher, C.C., Whelan, J., Daly, L. and Fitzpatrick, P., 2012. Socio-demographic, environmental, lifestyle and psychosocial factors predict self rated health in Irish Travellers, a minority nomadic population. Health & place, [online] 18(2), pp.330–338. available at: <10.1016/j.healthplace.2011.10.009> [accessed 15/07/2023]

Krzyzanowska, M.K., Kaplan, R. and Sullivan, R., 2011. How may clinical research improve healthcare outcomes? Annals of oncology, [online] 122, pp.vii10–vii15. available at: <10.1093/annonc/mdr420> [accessed 10/05/2023]

Lang, S.J., Abel, G.A., Mant, J. and Mullis, R., 2016. Impact of socioeconomic deprivation on screening for cardiovascular disease risk in a primary prevention population: a cross-sectional study. BMJ open, [online] 6(3), p.e009984. available at: <10.1136/bmjopen-2015-009984> [accessed 11/08/2023]

Marmot M, Allen J, Boyce T, Goldblatt P, Morrison J.(2020). Health Equity in England: The Marmot Review 10 Years On.1Institute of Health Equity; 2020 [online] available at: <https://www.instituteofhealthequity.org/resources-reports/marmot-review-10-years-on/the-marmot-review-10-years-on-executive-summary.pdf> [accessed 24/07/2023]

Moola, S., Munn, Z., Sears, K., Sfetcu, R., Currie, M., Lisy, K., Tufanaru, C., Qureshi, R., Mattis, P. and Mu, P., (2015). Conducting systematic reviews of association (etiology): The Joanna Briggs Institute’s approach. JBI Evidence Implementation, [online] 13(3), pp.163–169). available at: <https://journals.lww.com/ijebh/toc/2015/09000> [accessed 20/07/2023]

Munn, Z., Tufanaru, C. and Aromataris, E., (2014). JBI’s systematic reviews: data extraction and synthesis. AJN The American Journal of Nursing, [online] 114(7), pp.49–54. available at: <10.1097/01.naj.0000451683.66447.89> [accessed 15/07/2023]

Munn, Z., Stern, C., Aromataris, E., Lockwood, C. and Jordan, Z., (2018). What kind of systematic review should I conduct? A proposed typology and guidance for systematic reviewers in the medical and health sciences. BMC medical research methodology, [online] 18(1), pp.1–9. available at: 10.1186/s12874-017-0468-4 [accessed 15/07/2023]

Nanjo, A., Evans, H., Direk, K., Hayward, A.C., Story, A. and Banerjee, A., (2020). Prevalence, incidence, and outcomes across cardiovascular diseases in homeless individuals using national linked electronic health records. European Heart Journal, [online] 41(41), pp.4011–4020. available at: <10.1093/eurheartj/ehaa795> [accessed 19/04/2023]

NIHR (2020) Improving inclusion of under-served groups in clinical research: Guidance from the NIHR-INCLUDE project. UK: NIHR. [online] Available at: <https://www.nihr.ac.uk/documents/improving-inclusion-of-under-served-groups-in-clinical-research-guidance-from-include-project/25435> [accessed 25/07/2023]

NIHR (2021), Best Research for Best Health: The Next Chapter Our Operational Priorities [online] available at: <https://www.nihr.ac.uk/documents/about-us/best-research-for-best-health-the-next-chapter.pdf> [accessed 16/04/2023]

NIHR (2022a), Equality, Diversity & Inclusion Strategy 2022–2027 [online] available at: <https://www.nihr.ac.uk/documents/about-us/NIHR-equality-diversity-inclusion-strategy.pdf> [accessed 16/04/2023]

NIHR (2022b) Research ready communities and research champion’s pilot. [online] available at: <https://www.nihr.ac.uk/news/nihr-pilot-works-to-improve-awareness-and-inclusion-in-research/32008> [accessed 25/07/2023]

NIHR (2023a) Equality, Diversity and Inclusion Strategy, version 2.0 [online] available at: <https://www.nihr.ac.uk/documents/equality-diversity-and-inclusion-strategy-2022-2027/31295#overall-objectives> [accessed: 24/07/2023]

NIHR (2023b) Policy Research Programme - Guidance for Stage 1 Applications [online] available at: <https://www.nihr.ac.uk/documents/policy-research-programme-guidance-for-stage-1-applications-updated/26398?pr=> [accessed 17/07/2023]

Ojo-Fati, O., Joseph, A.M., Ig-Izevbekhai, J., Thomas, J.L., Everson-Rose, S.A., Pratt, R., Raymond, N., Cooney, N.L., Luo, X. and Okuyemi, K.S., (2017). Practical issues regarding implementing a randomized clinical trial in a homeless population: strategies and lessons learned. Trials, [online] 18, pp.1–10. available at: <10.1186/s13063-017-2046-9> [accessed 14/04/2023]

ONS (2014). What Does the 2011 Census Tell us about the Characteristics of Gypsy or Irish Travellers in England and Wales?. Office for National Statistics ONS [online] Available at: <https://www.ons.gov.uk/peoplepopulationandcommunity/culturalidentity/ethnicity/datasets/2011censusanalysiswhatdoesthe2011censustellusaboutthecharacteristicsofgypsyoririshtravellersinenglandandwales> [accessed: 14/04/2023]

ONS (2022a) Cost of living and depression in adults, Great Britain: 29 September to 23 October 2022 [online] available at: <https://www.ons.gov.uk/peoplepopulationandcommunity/healthandsocialcare/mentalhealth/articles/costoflivinganddepressioninadultsgreatbritain/29septemberto23october2022#:~:text=1.-,Main%20points,%2Dpandemic%20levels%20(10%25)> [accessed 15/07/2023]

ONS (2022b) Socioeconomic inequalities in avoidable mortality in England: 2020 [online] available at: <https://www.ons.gov.uk/peoplepopulationandcommunity/birthsdeathsandmarriages/deaths/bulletins/socioeconomicinequalitiesinavoidablemortalityinengland/2020#socioeconomic-inequalities-in-avoidable-mortality-by-cause> [accessed on 23/06/2023]

O’Reilly-de Brún, M., de Brún, T., Okonkwo, E., Bonsenge-Bokanga, J.S., De Almeida Silva, M.M., Ogbebor, F., Mierzejewska, A., Nnadi, L., van Weel-Baumgarten, E., van Weel, C. and van den Muijsenbergh, M., 2015. Using Participatory Learning & Action research to access and engage with ‘hard to reach’migrants in primary healthcare research. BMC Health Services Research, [online] 16(1), pp.1–16. available at: <10.1186/s12913-015-1247-8> [accessed 17/05/2023]

Page, M.J., Moher, D., Bossuyt, P.M., Boutron, I., Hoffmann, T.C., Mulrow, C.D., Shamseer, L., Tetzlaff, J.M., Akl, E.A., Brennan, S.E. and Chou, R., (2021). PRISMA 2020 explanation and elaboration: updated guidance and exemplars for reporting systematic reviews. bmj,1372. [online] available at: <https://www.bmj.com/content/372/bmj.n160> [accessed 27/07/2023]

Parry, G., Van Cleemput, P., Peters, J., Walters, S., Thomas, K. and Cooper, C., (2007). Health status of Gypsies and Travellers in England. Journal of Epidemiology & Community Health, [online] 61(3), pp.198–204. available at: <10.1136/jech.2006.045997> [accessed 25/04/2023]

Pearson, A., White, H., Bath-Hextall, F., Salmond, S., Apostolo, J. and Kirkpatrick, P., (2015). A mixed-methods approach to systematic reviews. JBI Evidence Implementation, [online] 13(3), pp.121–131. available at: <10.1097/xeb.0000000000000052> [accessed 15/07/2023]

Peters, J., Parry, G.D., Van Cleemput, P., Moore, J., Cooper, C.L. and Walters, S.J., 2009. Health and use of health services: a comparison between Gypsies and Travellers and other ethnic groups. Ethnicity & health, [online] 14(4), pp.359–377. available at: <10.1080/13557850802699130> [accessed 14/04/2023]

Popay, J., Roberts, H., Sowden, A., Petticrew, M., Arai, L., Rodgers, M., Britten, N., Roen, K. and Duffy, S., 2006. Guidance on the conduct of narrative synthesis in systematic reviews. A product from the ESRC methods programme Version, 1(1), [online] p.b92. available at: <https://citeseerx.ist.psu.edu/document?repid=rep1&type=pdf&doi=ed8b23836338f6fdea0cc55e161b0fc5805f9e27> [accessed 20/05/2023]

Public Health England (2019) Health Matters: Preventing Cardiovascular Disease [online] available at: <https://www.gov.uk/government/publications/health-matters-preventing-cardiovascular-disease/health-matters-preventing-cardiovascular-disease> [accessed 20/05/2023]

Public Health England, 2021. Community champions: A rapid scoping review of community champion approaches for the pandemic response and recovery. [online] available at: <https://assets.publishing.service.gov.uk/government/uploads/system/uploads/attachment_data/file/1011854/A_rapid_scoping_review_of_community_champion_approaches_for_the_pandemic_response_and_recovery_V8.pdf> [accessed 02/08/2023]

Quaife, S.L., Ruparel, M., Dickson, J.L., Beeken, R.J., McEwen, A., Baldwin, D.R., Bhowmik, A., Navani, N., Sennett, K., Duffy, S.W. and Wardle, J., 2020. Lung screen uptake trial (LSUT): randomized controlled clinical trial testing targeted invitation materials. American Journal of Respiratory and Critical Care Medicine, [online] 201(8), pp.965–975. available at: <10.1164/rccm.201905-0946OC> [accessed 17/05/2023]

Remes, O., Lafortune, L., Wainwright, N., Surtees, P., Khaw, K.T. and Brayne, C., (2019). Association between area deprivation and major depressive disorder in British men and women: a cohort study. BMJ open, [online] 9(11), p.e027530. available at: <10.1136/bmjopen-2018-027530> [accessed 20/06/2023]

Resnik, D.B., (2015). Bioethical issues in providing financial incentives to research participants. Medicolegal and bioethics, [online] pp.35–41. available at: <https://www.tandfonline.com/doi/full/10.2147/MB.S70416> [accessed 17/07/2023]

Rogers, A., Flynn, R.W., Mackenzie, I.S. and MacDonald, T.M., 2019. Does the provision of a DVD-based audio-visual presentation improve recruitment in a clinical trial? A randomised trial of DVD trial invitations. BMC Medical Research Methodology, [online] 19(1), pp.1–6. available at: <10.1186/s12874-019-0663-6> [accessed 17/05/2023]

Ryan R (2013) Cochrane Consumers and Communication Review Group. ‘Cochrane Consumers and Communication Review Group: data synthesis and analysis’. [online] available at: <https://cccrg.cochrane.org/sites/cccrg.cochrane.org/files/public/uploads/Analysis.pdf>[accessed 27/07/2023]

Sarkis-Onofre, R., Catalá-López, F., Aromataris, E. and Lockwood, C., (2021). How to properly use the PRISMA Statement. Systematic Reviews, [online] 10(1), pp.1–3. available at: <10.1186/s13643-021-01671-z> [accessed 20/07/2023]

Sethi, S., Kumar, A., Mandal, A., Shaikh, M., Hall, C.A., Kirk, J.M., Moss, P., Brookes, M.J. and Basu, S., (2021). The UPTAKE study: implications for the future of COVID-19 vaccination trial recruitment in UK and beyond. Trials, [online] 22, pp.1–12. available at: <https://link.springer.com/article/10.1186/s13063-021-05250-4> [accessed 20/07/2023]

Sharrocks, K., Spicer, J., Camidge, D.R. and Papa, S., (2014). The impact of socioeconomic status on access to cancer clinical trials. British journal of cancer, [online] 111(9), pp.1684–1687. available at <10.1038/bjc.2014.108> [accessed 19/06/2023]

Smith, D. and Ruston, A., (2013). ‘If you feel that nobody wants you you’ll withdraw into your own’: Gypsies/Travellers, networks and healthcare utilisation. Sociology of Health & Illness, [online] 35(8), pp.1196–1210. available at: <10.1111/1467-9566.12029> [accessed 15/07/2023]

Smith, L.T., (2021). Decolonizing methodologies: Research and indigenous peoples. Third Edition. London: Bloomsbury Publishing.

Smyth, L., McClements, L. and Murphy, P., 2020. Engaging hard-to-reach populations in research on health in pregnancy: the value of Boal’s simultaneous dramaturgy. Arts & Health, [online] 12(1), pp.71–79. available at: <10.1080/17533015.2018.1555176> [accessed 17/05/2023]

Snyder, H., (2019). Literature review as a research methodology: An overview and guidelines. Journal of business research, [online] 104, pp.333–339. available at: <10.1016/j.jbusres.2019.07.039> [accessed 20/07/2023]

Stafford, M., Steventon, A., Thorlby, R., Fisher, R., Turton, C. and Deeny, S., (2018). Briefing: Understanding the health care needs of people with multiple health conditions. London: Health Foundation. [online] available at: <https://www.health.org.uk/sites/default/files/upload/publications/2018/Understanding%20the%20health%20care%20needs%20of%20people%20with%20multiple%20health%20conditions.pdf> [accessed 19/06/2023]

Stern, C., Lizarondo, L., Carrier, J., Godfrey, C., Rieger, K., Salmond, S., Apostolo, J., Kirkpatrick, P. and Loveday, H., (2021). Methodological guidance for the conduct of mixed methods systematic reviews. JBI Evidence Implementation, [online] 19(2), pp.120–129. available at: <https://pubmed.ncbi.nlm.nih.gov/34061049/> [accessed 15/07/2023]

Sydor, A., (2013). Conducting research into hidden or hard-to-reach populations.1Nurse researcher, [online] 20(3). available at: <https://www.proquest.com/docview/1285579158/fulltextPDF/454E10ADEF97436FPQ/1?accountid=47912> [accessed 17/07/2023]

Taskforce on Multiple Condition, (2021). Evidence Review: Health equity and multiple long-term conditions. Richmond Group of Charities. [online] available at: <https://richmondgroupofcharities.org.uk/sites/default/files/evidence_review_final_22.07.pdf> [accessed 19/06/2023]

Tsamakis, K., Gadelrab, R., Wilson, M., Bonnici-Mallia, A.M., Hussain, L., Perera, G., Rizos, E., Das-Munshi, J., Stewart, R. and Mueller, C., (2021). Dementia in people from ethnic minority backgrounds: Disability, functioning, and pharmacotherapy at the time of diagnosis. Journal of the American Medical Directors Association, [online] 22(2), pp.446–452. available at <10.1016/j.jamda.2020.06.026> [accessed 10/05/2023]

Tully, L., Spyreli, E., Allen-Walker, V., Matvienko-Sikar, K., McHugh, S., Woodside, J., McKinley, M.C., Kearney, P.M., Dean, M., Hayes, C. and Heary, C., 2021. Recruiting ‘hard to reach’parents for health promotion research: experiences from a qualitative study. BMC Research Notes, [online] 14, pp.1–7. available at: <10.1186/s13104-021-05653-1> [accessed 17/05/2023]

Van Cleemput, P., Parry, G., Thomas, K., Peters, J. and Cooper, C., (2007). Health-related beliefs and experiences of Gypsies and Travellers: a qualitative study. Journal of epidemiology & community health, [online] 61(3), pp.205–210. available at: <10.1136/jech.2006.046078> [accessed 15/07/2023]

Van Cleemput, P., (2018). Health needs of Gypsy travellers. InnovAiT, [online] 11(12), pp.681–688. Available at: <10.1177/1755738018774075> [accessed 14/04/2023]

Van Cleemput, P., (2012). Providing healthcare to Gypsy and Traveller communities. Nursing in Practice, [online] 66(1), pp.26–8. available at: <https://www.researchgate.net/profile/Patrice-Van-Cleemput/publication/230719944_Providing_healthcare_to_Gypsy_and_Traveller_communities/links/0912f50365b2d214f5000000/Providing-healthcare-to-Gypsy-and-Traveller-communities.pdf> [accessed 15/07/2023]

Vickneson, K., Rogers, A., Anbarasan, T., Rorie, D.A., MacDonald, T.M. and Mackenzie, I.S., 2022. Factors influencing participation and long-term commitment to self-monitoring of blood pressure in a large remote clinical trial: The treatment in morning versus evening (TIME) study. Journal of Human Hypertension, [online] 36(12), pp.1099–1105. available at: <https://rdcu.be/djhCU> [accessed 17/05/2023]

Witham, M.D., Anderson, E., Carroll, C.B., Dark, P.M., Down, K., Hall, A.S., Knee, J., Maher, E.R., Maier, R.H., Mountain, G.A. and Nestor, G., (2020a). Ensuring that COVID-19 research is inclusive: guidance from the NIHR INCLUDE project. BMJ open, [online] 10(11), p.e043634. available at: https://bmjopen.bmj.com/content/bmjopen/10/11/e043634.full.pdf [accessed 20/07/2023]

Witham, M.D., Anderson, E., Carroll, C., Dark, P.M., Down, K., Hall, A.S., Knee, J., Maier, R.H., Mountain, G.A., Nestor, G. and Oliva, L., (2020b). Developing a roadmap to improve trial delivery for under-served groups: results from a UK multi-stakeholder process. Trials, [online] 21, pp.1–9. available at: <10.1186/s13063-020-04613-7> [accessed 20/04/2023]

